# Associations between fecal contamination of the household environment and enteric pathogen detection in children living in Maputo, Mozambique

**DOI:** 10.1101/2025.03.11.25323794

**Authors:** David A. Holcomb, Jackie Knee, Zaida Adriano, Drew Capone, Oliver Cumming, Erin Kowalsky, Rassul Nalá, Edna Viegas, Jill R. Stewart, Joe Brown

## Abstract

Environmental exposure to enteric pathogens is generally assessed using fecal indicators but relationships between markers of fecal contamination and actual exposure to enteric pathogens remain poorly characterized. We investigated whether *Escherichia coli* and two human fecal markers (HF183 and Mnif) in urban Mozambican household soil and drinking water were associated with detection in child stool of eight bacteria, three viruses, and three protozoa measured by multiplex reverse-transcription PCR and soil transmitted helminths assessed by microscopy. We used mixed-effects logistic regression with marginal standardization to obtain a pooled estimate of the overall indicator-pathogen relationship while simultaneously estimating pathogen-specific associations that accounted for assessing multiple pathogens per sample. At least one pathogen was detected in 88% (169/192) of child stools. Increasing drinking water *E. coli* gene concentration was associated with higher *Ascaris* prevalence, while human HF183 in drinking water was weakly associated with lower prevalence of the most common pathogens but was infrequently detected. No fecal marker in soil was clearly associated with any pathogen. We did not find evidence to support human markers as reliable indicators of enteric pathogen carriage in a high-prevalence domestic setting and recommend targeting enteric pathogens directly.

**Table of Contents Graphic:** 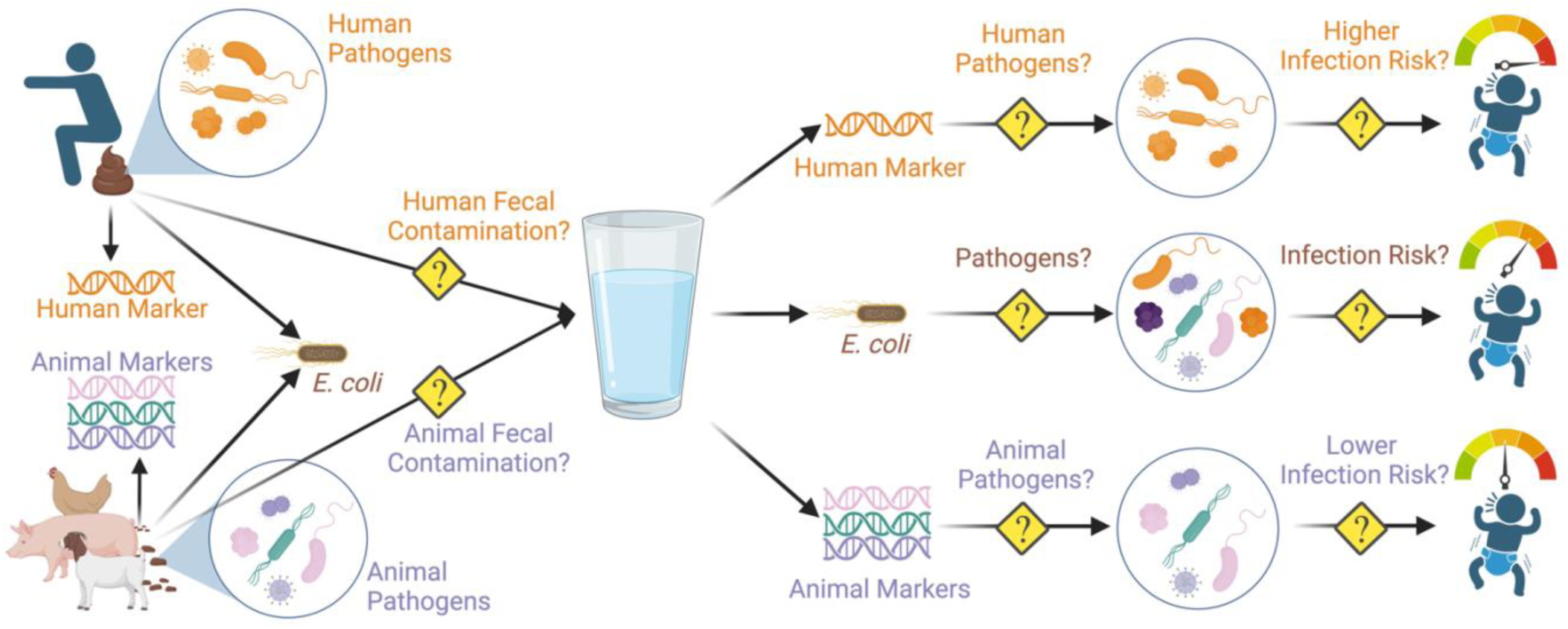

## Introduction

Children living in settings with inadequate water, sanitation, and hygiene (WASH) are exposed to numerous enteric pathogens early in life.^1,2^ Several recent, large-scale interventions to improve WASH conditions did not consistently improve child health outcomes in robust evaluation studies, despite high intervention fidelity and adherence.^3–10^ It has been suggested that failure of the interventions to adequately interrupt environmentally mediated pathogen transmission and prevent exposure to enteric pathogens might account for the limited improvements to child health.^11–13^ Determining which environmental compartments are contaminated with and mediate exposure to enteric pathogens could inform improved design and targeting of interventions to prevent fecal-oral transmission of disease.

Detecting an enteric pathogen in stool unambiguously indicates that the individual was previously exposed to that pathogen.^14^ Assessing enteric pathogens is challenging using traditional diagnostic approaches owing to the diversity of potential agents, but modern molecular microbial detection techniques enable the simultaneous detection of multiple enteric pathogens in a single sample.^15^ The typically low abundance of pathogens in environmental compartments has historically made direct pathogen detection in environmental samples particularly unreliable. Instead, more abundant and easily detected fecal indicator bacteria like *Escherichia coli*, which are shed in the feces of humans and other warm-blooded animals, have traditionally been used as markers of fecal contamination to infer the likely presence of enteric pathogens.^16^ Host-associated fecal markers have increasingly been used to determine the source of fecal contamination; because many pathogens are host-specific, it has been suggested that such host-associated markers (particularly those indicating human fecal contamination) might also be more strongly associated with risks to human health.^17^

The aim of this study was to investigate whether fecal markers in household environments were reliable indicators of child exposure to enteric pathogens. We assessed markers of general and human-specific fecal contamination in water and soil and compared them with enteric pathogens detected in child stools from low-income households in urban Maputo, Mozambique.^18^ We implemented a hierarchical modeling approach to simultaneously estimate fecal marker associations with the prevalence of each individual pathogen and pooled across all pathogens to account for the multiple pathogens assessed in each stool sample.^19^

## Materials and Methods

### Study Setting and Design

We conducted this analysis as a nested sub-study of the Maputo Sanitation (MapSan) trial, which evaluated the impact of an onsite sanitation intervention on child enteric infections in 16 densely populated, low-income, low-income neighborhoods of urban Maputo, Mozambique.^7,18^ The intervention was implemented between February 2015 and February 2016 and replaced shared sanitation facilities in poor condition in compounds (household clusters sharing sanitation and outdoor living space).^20^ Intervention compounds were selected by the implementing organization, Water and Sanitation for the Urban Poor (WSUP), according to previously described engineering and demand criteria.^7,21^ Compounds with similarly poor-condition shared sanitation, numbers of residents, and a legal piped water supply that expressed willingness to contribute financially to sanitation facility construction costs but had not received the intervention were selected to serve as controls.

The parent MapSan trial assessed enteric pathogen and diarrhea outcomes for an open cohort of children living in study compounds in two cross-sectional surveys conducted approximately one year apart.^18^ Baseline (pre-intervention) visits to intervention compounds were conducted approximately two weeks before the intervention facilities were available for use. Baseline visits to control compounds were conducted concurrently to limit the potential impacts of secular trends and seasonality. Follow-up visits to intervention compounds were conducted 12 months (± 2 weeks) after residents began using the intervention facilities and control compounds were concurrently revisited. All children aged 1-48 months living in the study compound were eligible for enrollment during the baseline visit. At the 12-month follow-up visit, all children who were or would have been eligible during the baseline visit and who had been living in the compound for at least 6 months (or since birth, if younger than 6 months) were eligible. For a subset of compounds during baseline enrollment in May – August 2015 and again during the 12-month follow-up in June – September 2016, we concurrently sampled environmental matrices from all households with children participating in the MapSan study.^21^ We conducted the nested sub-study of fecal marker associations with diarrhea and detection of enteric pathogens for this subset of compounds from which we both enrolled children in the parent trial and collected environmental samples.

### Household Visit Procedures

Trained enumerators first obtained verbal consent from the compound head to conduct study activities before seeking written, informed consent from the parent or guardian of each eligible child. Activities were conducted in Portuguese or Changana, as preferred by the participant, and written materials were provide in Portuguese. After completing consent procedures, enumerators conducted compound-, household-, and child-level surveys; measured child weight and height; and provided caregivers with materials to collect stool samples.^7,22^ Compounds were revisited the following day to retrieve child stools and collect environmental samples. We attempted to collect a stool from each enrolled child; if unsuccessful after multiple attempts, a registered nurse collected a rectal swab with the written consent of a parent or guardian. After completing all sample collection activities, representatives of the National Deworming Campaign at the Mozambican Ministry of Health offered single-dose albendazole (400 mg; 200 mg for children aged 6–12 months) to all non-pregnant compound members older than 6 months.

While retrieving child stool samples, we collected stored drinking water and soil at the entrance of each household with a child enrolled in the study, as well as soil at the entrance to the compound latrine.^21^ As described previously, caregivers were asked to provide ∼1 L of household stored water as if they were giving a child water to drink (i.e., from the same source and in the same drinking vessel that would be served to a child).^23^ Soil was collected 1 m outside of the latrine entrance or household entrance using a metal scoop sanitized with 10% bleach and 70% ethanol to homogenize a 10 cm × 10 cm area to a depth of ∼1 cm.^24,25^ Both stool and environmental samples were transported on ice to the Molecular Parasitology Laboratory at Instituto Nacional de Saúde (INS) in Maputo City within 6 hours of collection.

### Laboratory Analysis

Stool was analyzed for soil-transmitted helminths (STH) by INS technicians using the single-slide Kato-Katz microscopy method (Vestergaard Frandsen, Lausanne, Switzerland); *Ascaris lumbricoides* and *Trichuris trichiura* were the only STH that we routinely observed by the Kato-Katz method. The remaining sample was stored in 2 mL aliquots at -80 °C awaiting molecular analysis.^7,26^ We determined soil moisture content by drying approximately 5 g wet soil by microwave oven in 5-minute increments until the measured weight stabilized.^20,25,27^ We manually eluted 1 g of each soil sample in 100 mL of autoclaved, distilled water. Up to 1 mL of this soil eluate and 100 mL of stored water were processed by membrane filtration and cultured on mTEC broth (HiMedia Laboratories, Mumbai, India) to enumerate viable *E. coli*.^28^ We filtered an additional 300 mL of stored water and 30 mL of soil eluate for molecular analysis and immediately stored the filters at -80 °C.

Frozen samples were shipped on dry ice with temperature monitors to Georgia Institute of Technology (Atlanta, GA, USA), whereupon the environmental sample filters were transferred on dry ice to University of North Carolina (Chapel Hill, NC, USA). Detailed descriptions of the molecular analyses conducted for both the stool and the environmental samples have been published previously and are summarized in the Supporting Information (SI).^7,21–23^ Briefly, stool samples were analyzed by multiplex reverse-transcription polymerase chain reaction (RT-PCR) using the Luminex xTAG Gastrointestinal Pathogen Panel (GPP, Luminex Corp, Austin, TX) to detect 14 enteric pathogens:

### Campylobacter jejuni/coli/lari; Clostridioides difficile toxin A/B; enterotoxigenic E. coli (ETEC)

LT/ST; Shiga toxin-producing *E. coli* (STEC) *stx1*/*stx2*; *E. coli* O157; *Shigella* spp.; *Vibrio cholerae*; *Yersinia enterocolitica*; *Giardia lamblia*; *Cryptosporidium parvum/hominis*; *Entamoeba histolytica*; adenovirus 40/41; norovirus GI/GII; and rotavirus.^22,29^ We excluded the GPP *Salmonella* spp. target following multiple reports of poor assay specificity.^7,30,31^ Stool samples were treated with MS2 bacteriophage as a specimen processing control (SPC), nucleic acid extractions included a negative extraction control (NEC) consisting of only lysis buffer and MS2, and no-template controls (NTC) with molecular-grade water were included with each GPP analysis.

Soil and water sample filters were analyzed by locally validated quantitative (real-time) PCR (qPCR) to assess molecular markers of fecal contamiantion.^16,23,32^ We quantified the *E. coli* 23S gene (EC23S857) and two human-associated fecal markers, *Bacteroides dorei* 16S rRNA (HF183/BacR287) and *Methanobrevibacter smithii nifH* gene (Mnif).^33–35^ Filters were treated with salmon testes DNA prior to extraction as an SPC, a clean filter treated with salmon DNA was included in each DNA extraction batch as the NEC, and NTCs were included on each qPCR plate.

### Statistical Analysis

All children enrolled in MapSan during the baseline or 12-month follow-up phases with at least one concurrently collected stored water or soil sample were considered for this analysis. We conducted a cross-sectional analysis of all observations from both study phases, including multiple observations for children enrolled at both time points. Outcomes included the individual prevalence of each enteric pathogen detected in at least five samples and the seven-day period prevalence of caregiver-reported diarrhea, defined as passing at least three loose or watery stools or any bloody stool in a 24-hour period in the previous seven days.^18,36,37^ Diarrhea was ascertained via surveys conducted with caregivers immediately following enrollment, as were child sex and age, caregiver education, and household assets.^22^ We calculated asset-based household wealth scores using the Simple Poverty Scorecard for Mozambique.^38^

*E. coli* concentrations were expressed as colony forming units (cfu) or gene copies per 100 mL water or 1 dry gram of soil and were log_10_-transformed for all analyses. We considered samples in which *E. coli* colonies or genes were not detected to be censored below the limit of detection (LOD). *E. coli* log_10_ concentrations were imputed for censored samples as the expected value of a normal distribution truncated at the sample-specific process LOD (Table S4).^23,39^ The mean and standard deviation of the truncated normal distribution were obtained as the maximum likelihood estimates (MLE) of a censored normal distribution fit to the observed log_10_ *E. coli* concentrations for each assay and sample matrix. We imputed concentrations using the *truncnorm* R package with censored normal distribution MLEs obtained by the *fitdistrplus* package.^40,41^ The correspondence between *E. coli* colony count and gene copy concentrations was assessed by non-parametric Spearman’s rank correlation. Because human-associated markers were infrequently detected in these samples, we did not impute concentrations for HF183 or Mnif.^21^ We instead analyzed the binary presence/absence of HF183 and Mnif in stored water and soil samples.

We used mixed effects logistic regression models to estimate fecal marker associations with the prevalence of specific enteric pathogens in child stool and the prevalence of caregiver-reported diarrhea. Regression models were implemented using the approach described by Holcomb et al. (2023) to simultaneously estimate pathogen-specific associations and a pooled association across pathogens that represents the expected association for a “typical” pathogen.^19^ We estimated marginal prevalence ratios (PR) and prevalence differences (PD) as measures of association for a ten-fold (1 log_10_) increase in *E. coli* concentration or for the detection of a human-associated marker.^42^ Separate models were fit for each combination of marker and sample matrix and all models were adjusted for pre-selected covariates: child age, child sex, caregiver education (completion of primary school), and household wealth score.^7^ Samples missing covariate data were excluded. We did not consider sanitation-related variables because sanitation was presumed to be upstream of fecal contamination on the causal chain and our aim was to assess associations of fecal markers with pathogen carriage and illness.^43^

The detection status of each pathogen was treated as a separate observation of the same binary response variable, producing a long-format model design matrix with multiple rows per child stool (one row for each pathogen). We allowed the intercept and the slopes for all predictor variables (i.e., the fecal marker exposure variable and the child covariates) to vary by pathogen, with each pathogen-specific slope providing the predictor’s estimated association with that pathogen. The intercept was also allowed to vary by child stool to account for measuring multiple pathogens per sample and by compound to address clustering of multiple children within compounds. We treated all the pathogen-specific slopes for a given predictor as a group with a shared mean and variance.^44,45^ The group mean corresponded to the weighted-average association for that predictor across all pathogens and the group variance indicated the extent to which the association differed between pathogens. We also modeled the covariance between the pathogen-varying intercept and slopes to account for dependencies between predictors. By partially pooling information across pathogens, this model structure helps to regularize the estimated associations for each individual pathogen, thereby limiting the potential for inflated false discovery rates from conducting multiple comparisons.^44,46,47^ The full model equation and prior distribution selections are presented in the SI.

We used the *brms* package in R version 4.2.2 to specify models and conduct Markov chain Monte Carlo (MCMC) via the *cmdstanr* backend.^45,48–50^ Models were fit using the default no-U-turn sampler with four chains of 1000 warm-up and 1000 sampling iterations each, for 4000 total posterior samples. We specified weakly informative prior distributions (described in the SI) to regularize parameter estimates and facilitate computation.^21,44,51^ We implemented marginal standardization to recover estimates of the associations between fecal marker exposure and the prevalence of each pathogen in stool.^42^ For the binary human marker exposures, we used the fitted models to predict the probability of detecting each pathogen under two scenarios: none of the children were exposed to the fecal marker and all children were exposed. All other child covariate values remained unchanged, such that the predicted pathogen probabilities across all stools retained the distribution of confounders in the sample population. An analogous approach was adopted for the continuous *E. coli* exposures using finite differences to numerically approximate the slope corresponding to a log_10_ increase in *E. coli* concentration (see SI).^52,53^ We sampled predicted probabilities for each observation in the model from the 4000 posterior samples using the *tidybayes* package.^54–56^ The posterior predictive distribution of the prevalence ratio was obtained by dividing the exposed predicted probabilities by their corresponding unexposed predicted probabilities. Likewise, subtracting the unexposed predicted probabilities from the exposed probabilities provided the posterior predictive distribution of the prevalence difference.^57^ We summarized the marginal PR and PD estimates for each pathogen as the mean and central 95% probability interval (95% CI) of the corresponding posterior predictive distribution. PR and PD estimates with 95% CIs that excluded the null value (one for PR, zero for PD) were considered significant.

Fecal marker associations with diarrhea prevalence were estimated using the same approach after removing all pathogen-varying and stool-varying terms from the model. We also estimated associations with the four pathogen classes (bacteria, viruses, protozoa, and STH) by replacing the individual pathogens in the model with binary composite-class outcomes that represented detection of one or more pathogens of that class.^7^

### Ethical Approval and Data Availability

This study was approved by the Comité Nacional de Bioética para a Saúde, Ministério da Saúde, Republic of Mozambique (333/CNBS/14), the Research Ethics Committee of the London School of Hygiene & Tropical Medicine (reference 8345), the Institutional Review Board of the University of North Carolina at Chapel Hill (IRB 15-0963), and the Institutional Review Board of the Georgia Institute of Technology (protocol H15160). The parent trial was pre-registered at ClinicalTrials.gov (NCT02362932). The deidentified participant-level data and R code used in this analysis are available at https://daholcomb.github.io/manuscripts/mapsan_mst/.

## Results

### Participant and Sample Characteristics

We obtained 194 participant observations with at least one matching environmental sample and complete covariate data for child sex and age, caregiver education, and household wealth. Observations represented 156 unique participants, 38 of whom were enrolled in both survey rounds. Most observations included both caregiver-reported diarrhea status (n=193) and stool samples analyzed for enteric pathogens by GPP (n=192); fewer stools (n=169) were analyzed by microscopy for STH. The proportions of observations from female participants, each age group, and caregivers who completed primary school were similar across the three outcome types, as was the median household wealth score (Table 1). Participant observations were matched to 154 stored water samples and 233 soil samples. Multiple soil samples were collected from each compound and each environmental sample was separately matched to each participant observation from the corresponding household/compound, resulting in 194 participant observation/water sample pairs and 354 participant observation/soil sample pairs.

**Table 1.**
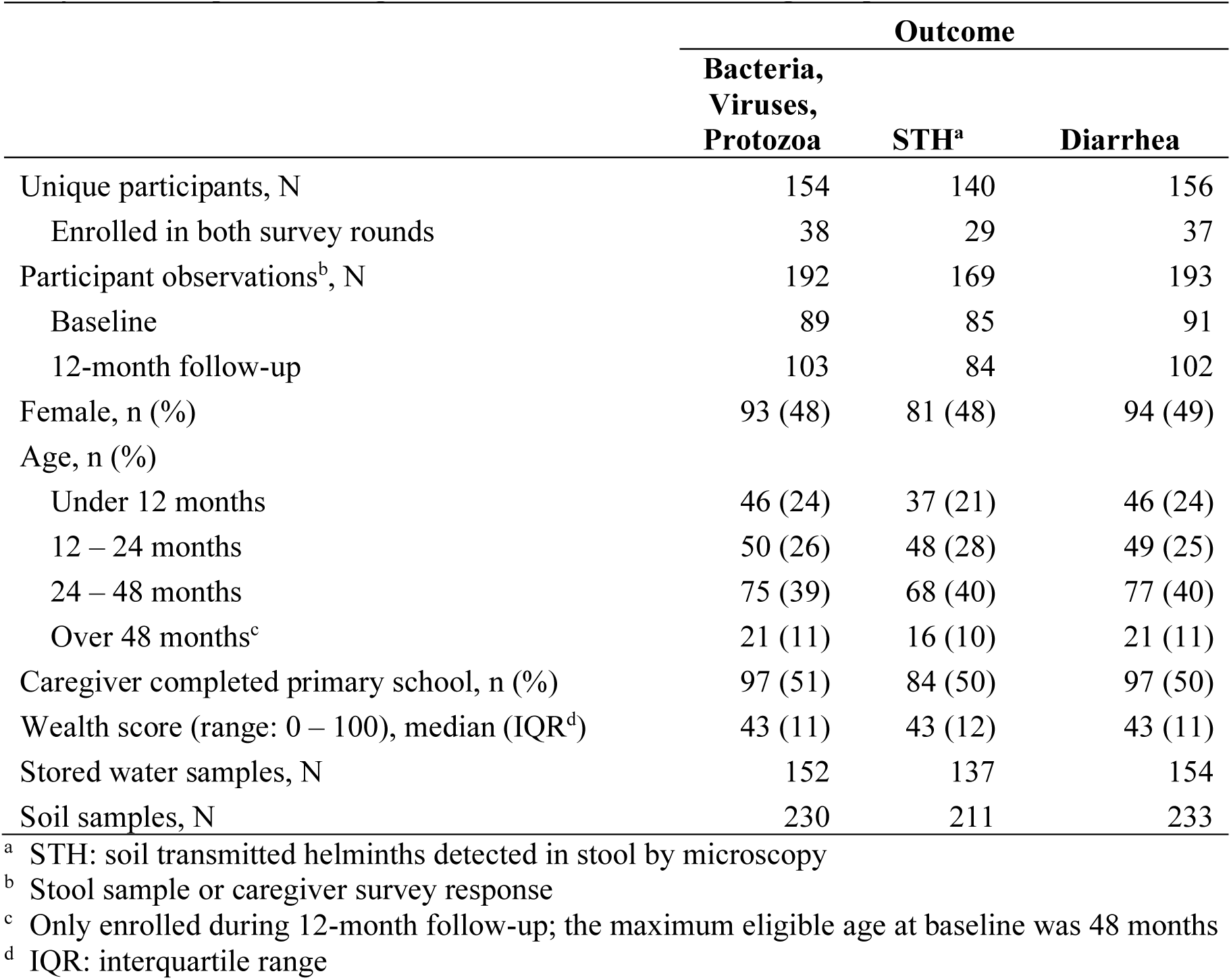
Participant demographics and sample characteristics by outcome type included in MapSan sub-study evaluating relationships between fecal markers and pathogen.

|  | Outcome |  |  |
| --- | --- | --- | --- |
|  | Bacteria,<br>Viruses,<br>Protozoa | STH<sup>a</sup> | Diarrhea |
| Unique participants, N | 154 | 140 | 156 |
| Enrolled in both survey rounds | 38 | 29 | 37 |
| Participant observations<sup>b</sup>, N | 192 | 169 | 193 |
| Baseline | 89 | 85 | 91 |
| 12-month follow-up | 103 | 84 | 102 |
| Female, n (%) | 93 (48) | 81 (48) | 94 (49) |
| Age, n (%) |  |  |  |
| Under 12 months | 46 (24) | 37 (21) | 46 (24) |
| 12 – 24 months | 50 (26) | 48 (28) | 49 (25) |
| 24 – 48 months | 75 (39) | 68 (40) | 77 (40) |
| Over 48 months<sup>c</sup> | 21 (11) | 16 (10) | 21 (11) |
| Caregiver completed primary school, n (%) | 97 (51) | 84 (50) | 97 (50) |
| Wealth score (range: 0 – 100), median (IQR<sup>d</sup>) | 43 (11) | 43 (12) | 43 (11) |
| Stored water samples, N | 152 | 137 | 154 |
| Soil samples, N | 230 | 211 | 233 |

<sup>a</sup> STH: soil transmitted helminths detected in stool by microscopy
<sup>b</sup> Stool sample or caregiver survey response
<sup>c</sup> Only enrolled during 12-month follow-up; the maximum eligible age at baseline was 48 months
<sup>d</sup> IQR: interquartile range

### Enteric Pathogen Detection and Diarrhea Outcomes

We detected at least one enteric pathogen in 169 of the 192 stools (88%) with paired environmental samples. Pathogenic bacteria, detected in 133 (69%) stools, were the most common pathogen class (Table 2), and *Shigella* was the highest prevalence individual pathogen (n=100, 52%). Protozoa were detected in 103 stools (54%), with *Giardia* (n=96, 50%) accounting for nearly all protozoan detections. STH were detected almost as frequently (n=85, 50% of the 169 stools tested for STH) and *Trichuris* (n=75, 44%) accounted for the majority of STH detections. The lowest prevalence pathogen class was viruses, which were detected in only 30 stools (16%). Norovirus GI/GII (n=27, 14%) was the only virus detected more than twice. Caregivers reported diarrhea symptoms in the preceding week for 19 participant observations (10%). Enteric pathogen prevalence was similarly high in the full MapSan cohort, although diarrhea prevalence was slightly lower in this sub-study (10%) than in the MapSan parent study (13%).^7,22^

**Table 2.** Enteric pathogen and diarrhea outcomes observed in the MapSan sub-study.

| Outcome | N | Positive |  |
| --- | --- | --- | --- |
|  |  | n | % |
| Any bacteria | 192 | 133 | 69 |
| *C. difficile* |  | 10 | 5 |
| *Campylobacter* |  | 23 | 12 |
| *E. coli* O157 |  | 2 | 1 |
| ETEC |  | 51 | 27 |
| STEC |  | 8 | 4 |
| *Shigella* |  | 100 | 52 |
| *V. cholerae* |  | 0 | 0 |
| *Y. enterocolitica* |  | 0 | 0 |
| Any viruses | 192 | 30 | 16 |
| Adenovirus 40/41 |  | 2 | 1 |
| Norovirus GI/GII |  | 27 | 14 |
| Rotavirus |  | 1 | 0.5 |
| Any protozoa | 192 | 103 | 54 |
| *Cryptosporidium* |  | 7 | 4 |
| *E. histolytica* |  | 4 | 2 |
| *Giardia* |  | 96 | 50 |
| Any STH | 169 | 85 | 50 |
| *Ascaris* |  | 33 | 20 |
| *Trichuris* |  | 75 | 44 |
| Diarrhea | 193 | 19 | 10 |

### Fecal Contamination of the Domestic Environment

*E. coli* contamination of household stored water and domestic soil was widely detected by both culture and molecular approaches. Culturable *E. coli* was detected in 129 (85%) of the 152 water samples tested, with a mean concentration of 1.4 log_10_ cfu/100 mL. Of the 222 soil samples tested, 210 (95%) were culture-positive for *E. coli* at a mean concentration of 3.8 log_10_ cfu/dry g. *E. coli* gene targets were detected with slightly greater frequency, in 90% of stored water samples (136/152) and 98% of soil samples (228/233). *E. coli* colony counts and gene copy concentrations were positively correlated in both water (Spearman’s *ρ*: 0.40; *p* < 0.001) and soil (Spearman’s *ρ*: 0.47; *p* < 0.001) samples, although the molecular *E. coli* measurements were less variable than the culture-based measurements in water samples (Table 3). The moderate correlation magnitudes suggest that for any given sample, the culture and molecular measurements may indicate somewhat differing levels of *E. coli*, but both approaches broadly identified higher *E. coli* quantities in the same samples.

**Table 3.** Detection and concentration of fecal markers in water and soil samples by paired outcome type.

| Outcome type | Exposure matrix | Fecal marker | Observations |  |  | Fecal marker |  |
| --- | --- | --- | --- | --- | --- | --- | --- |
|  |  |  | Outcome N | Exposure N | Paired<sup>a</sup> N | Detected n (%) | log<sub>10</sub> concentration mean (SD<sup>b</sup>) |
| Bacteria, Viruses, Protozoa | Water | *E. coli* colonies | 185 | 150 | 185 | 161 (87) | 1.4 (1.4) |
|  |  | *E. coli* genes | 186 | 150 | 186 | 167 (90) | 4.2 (0.8) |
|  |  | HF183 | 186 | 150 | 186 | 30 (16) |  |
|  |  | Mnif | 184 | 149 | 184 | 1 (0.5) |  |
|  | Soil | *E. coli* colonies | 183 | 219 | 333 | 319 (96) | 3.8 (1.0) |
|  |  | *E. coli* genes | 189 | 230 | 349 | 344 (99) | 6.8 (1.0) |
|  |  | HF183 | 189 | 230 | 349 | 165 (47) |  |
|  |  | Mnif | 189 | 229 | 347 | 157 (45) |  |
| STH | Water | *E. coli* colonies | 163 | 136 | 163 | 143 (88) | 1.5 (1.4) |
|  |  | *E. coli* genes | 163 | 135 | 163 | 146 (90) | 4.3 (0.8) |
|  |  | HF183 | 163 | 135 | 163 | 28 (17) |  |
|  |  | Mnif | 161 | 134 | 161 | 1 (0.5) |  |
|  | Soil | *E. coli* colonies | 161 | 200 | 295 | 283 (96) | 3.8 (1.0) |
|  |  | *E. coli* genes | 167 | 211 | 311 | 306 (98) | 6.8 (1.0) |
|  |  | HF183 | 167 | 211 | 311 | 146 (47) |  |
|  |  | Mnif | 167 | 210 | 310 | 142 (46) |  |
| Diarrhea | Water | *E. coli* colonies | 186 | 152 | 186 | 162 (87) | 1.4 (1.4) |
|  |  | *E. coli* genes | 187 | 152 | 187 | 168 (90) | 4.3 (0.8) |
|  |  | HF183 | 187 | 152 | 187 | 30 (16) |  |
|  |  | Mnif | 185 | 151 | 185 | 1 (0.5) |  |
|  | Soil | *E. coli* colonies | 184 | 222 | 335 | 321 (96) | 3.8 (1.0) |
|  |  | *E. coli* genes | 190 | 233 | 351 | 346 (99) | 6.8 (1.0) |
|  |  | HF183 | 190 | 233 | 351 | 164 (47) |  |
|  |  | Mnif | 190 | 232 | 349 | 157 (45) |  |

<sup>a</sup> Each environmental sample was paired with all participant observations from the same household/compound and each participant observation was paired with all environmental samples from the same household/compound; each unique participant observation and environmental sample pair was considered a separate observation
<sup>b</sup> SD: standard deviation

Human-associated markers were less common, particularly in stored water, for which Mnif was only detected in a single sample and HF183 in 23 samples (15%). The two human fecal markers were detected at similar frequencies in soil samples, 39% of which were HF183 positive (90/233) and 38% Mnif positive (87/232). At least one human marker was detected in 60% of soil samples (140/232), but the two markers were only co-detected in 36 samples—26% of the human marker-positive samples and 16% of all soil samples. Table 3 presents the fecal marker detection frequency and mean log_10_ concentration among all pairs of participant observations and environmental samples by outcome type. Because each participant observation could be paired with multiple environmental samples and each sample paired with multiple participant observations, the number of paired outcome/exposure observations exceeded the number of environmental samples collected and, for soil samples, also exceeded the number of participant observations. The varying number of participant observations obtained by each outcome ascertainment method (stool GPP, stool microscopy, and caregiver survey) also resulted in different numbers of paired observations for each outcome type. However, the corresponding variation in average fecal marker exposure values was minimal (Table 3).

### Fecal Marker Associations with Enteric Pathogen and Diarrhea Prevalence

Fecal markers in the domestic environment were not consistently associated with an increased prevalence of enteric pathogen detection or of reported diarrhea (Figure 1, Table S8). No individual pathogen was clearly associated with *E. coli* culture concentrations in household stored water or domestic soils. Likewise, *E. coli* gene concentrations in soil were not significantly associated with the prevalence any individual pathogen. By contrast, we observed a higher prevalence of every pathogen and of diarrhea with increasing *E. coli* gene concentrations in household stored water. In particular, the prevalence of *Ascaris* increased by 42% (95% CI: 6%, 100%) for every log_10_ increase in *E. coli* gene copies/100 mL in water. For all other outcomes, the 95% CIs for the PR and PD estimates (Figure S1) included the null, but the point estimates were consistently positive. Pooled across all pathogens, the prevalence of a typical pathogen was 20% higher (95% CI: -21%, 71%), on average, for every log_10_ increase in *E. coli* genes in stored water.

**Figure 1.**
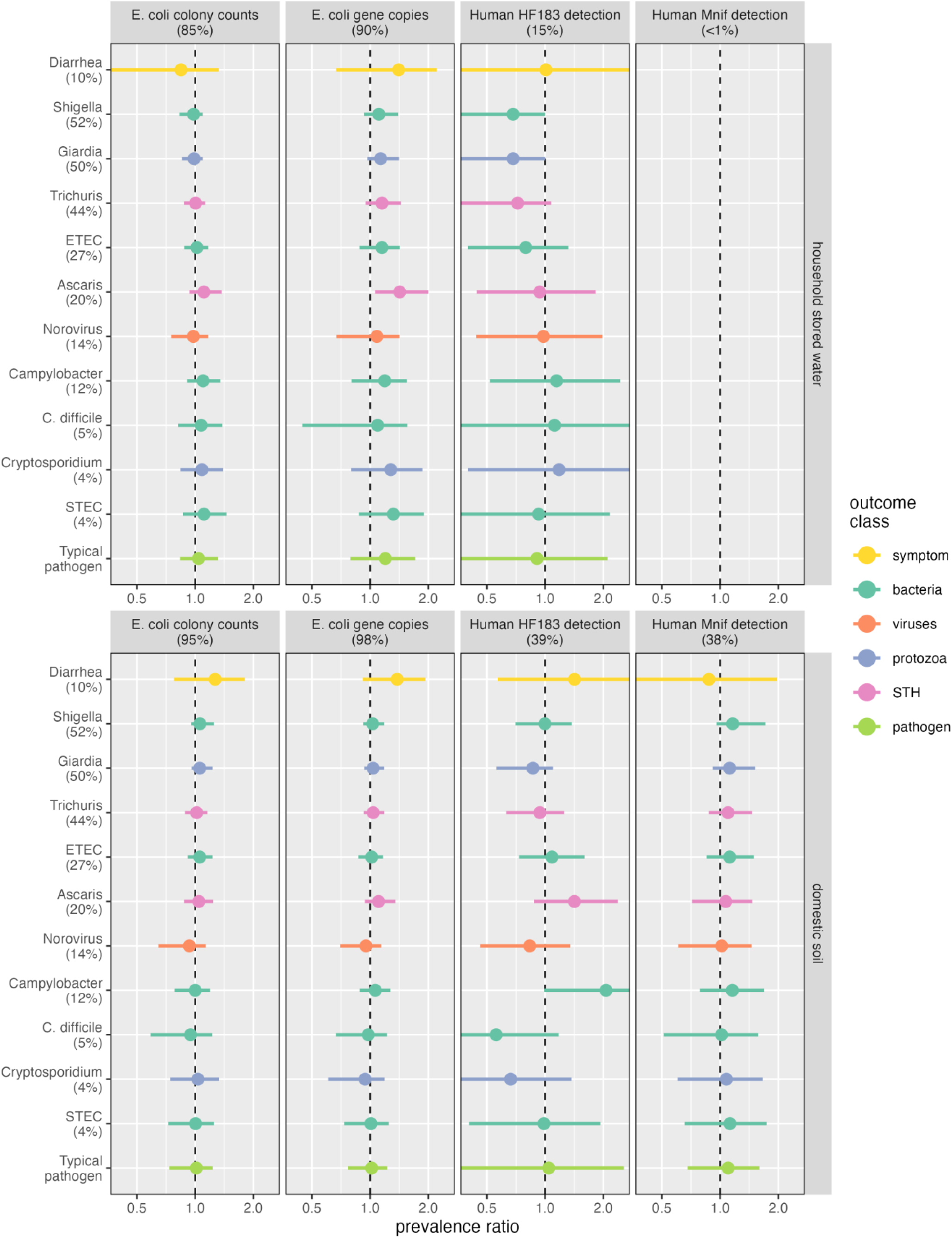
Mean and 95% CI marginal prevalence ratio estimates (horizontal axis) of diarrhea and of enteric pathogens in stool (vertical axis) for a ten-fold increase in *E. coli* concentration (left-two columns) or detection of a human fecal marker (right-two columns) in household stored water (top panel) and domestic soil (bottom panel). Percentages in parentheses correspond to the detection frequency for each fecal marker and outcome, with color indicating the outcome class.

Detection of the human fecal marker HF183 in stored water was associated with lower prevalence of the four most common pathogens. The inverse associations were strongest for the highest-prevalence pathogens, *Shigella* (PR: 0.68; 95% CI: 0.26, 1.00) and *Giardia* (PR: 0.68; 95% CI: 0.29, 1.00), each detected in about half of all stools. Associations attenuated as prevalence decreased: the PR for *Trichuris*, present in 44% of stools, was 0.72 (95% CI: 0.35, 1.08) and ETEC, detected in 27% of stools, had a PR of 0.79 (95% CI: 0.40, 1.32). No pathogen that was detected in less than a quarter of stool samples was clearly associated with HF183 in stored water. Diarrhea, observed in only 10% of participants, was likewise unassociated with stored water HF183 detection. Pathogen associations with HF183 detection in soil were inconsistent. *Campylobacter* prevalence in stool was doubled for HF183-positive soil (PR: 2.06; 95% CI: 0.99, 4.13) but the estimate was imprecise and included the null.

*Ascaris* prevalence was also elevated, while *C. difficile* and *Cryptosporidium* prevalence was diminished, though none were statistically significant. The estimated pooled association of HF183 in soil across all pathogens was close to the null and imprecise (PR: 1.04; 95% CI: 0.31, 2.56). The PR point estimates for Mnif exposure in soil were slightly above the null for all pathogens (but not for diarrhea) and were generally more precise than the corresponding estimates for HF183 in soil. However, Mnif soil exposure associations were not significant for any pathogen and the pooled estimate across pathogens was most consistent with no association (PR: 1.10; 95% CI: 0.68, 1.72).

No pathogen class was significantly associated with *E. coli* concentration or human marker detection; across all four pathogen classes, the pooled PR was close to the null for all fecal markers in both water and soil (Figure S2). Patterns of association were comparable on both the absolute and relative scales, with the same positioning of PR and PD point estimates and 95% CIs relative to the null for all outcomes, fecal indicators, and sample matrices (Figure S1, Figure S3, Table S8).

The association of HF183 in household stored water with lower prevalence of the four most common pathogens in this study might be an artifact of the frequency with which these pathogens were detected relative to the scarcity of human marker detections in stored water. *Shigella*, *Giardia,* and *Trichuris*—representing three distinct pathogen classes—were each present in roughly half of stools (Table 2), while HF183 was detected in only 23 (15%) water samples, corresponding to 30 HF183-positive stool pathogen/water sample pairs out of 186 pairs total (for STH outcomes, 28 HF183-positive pairs of 163 total). Across every pathogen tested, the majority (20–28) of stool/water pairs where HF183 was detected was negative for the pathogen (Table S6). This pattern was consistent regardless of the number of pathogen-positive schools, resulting in higher prevalence pathogens having a larger proportion of positive stools paired with HF183-negative water samples. Though this corresponds to HF183 detection being associated with lower pathogen prevalence, the relationship appears to have been driven by how frequently the outcome was detected rather than by changes in the distribution of the exposure. The number of HF183-positive samples that paired with pathogen-negative stools did not increase when pathogens were inversely associated with HF183 in water. Instead, the number of stools positive for these pathogens increased while the number of HF183-positive, pathogen-negative pairs remained stable.

## Discussion

Host-associated fecal markers have successfully been used to identify sources of fecal pollution in environments including surface waters,^58,59^ ground water,^60,61^ and agricultural water sources,^62,63^ but their utility in highly contaminated domestic environments has been more limited.^16^ With some exceptions,^64^ human-associated fecal markers have demonstrated poor diagnostic accuracy in such settings, both failing to be detected in human feces and frequently cross-reacting with the other animal species that commonly co-inhabit domestic environments in resource-limited settings.^21,65–68^ A recent meta-analysis of household-level sanitation interventions found no effect on host-associated fecal markers despite observing modest reductions in enteric pathogens in the same domestic environments.^69^ It is notable that enteric pathogens provided a stronger signal of changes in fecal contamination when the difficulty of reliably ascertaining enteric pathogens in the environment motivated the use of fecal markers in the first place. A companion meta-analysis likewise found that environmental enteric pathogen detection was associated with subsequent child enteric infections and diarrhea, while host-associated fecal markers were not associated with any child health outcome.^70^

Traditional fecal indicator bacteria, particularly *E. coli*, have proved somewhat more reliable than host-associated markers for assessing the impacts of efforts to mitigate domestic fecal contamination and as indicators of child health risk. Only viable organisms are reflected in colony counts from *E. coli* culture, which might better correspond to hazards present from viable enteric pathogens than molecular fecal markers that can be detected from dead and non-viable cells. However, molecular signals from human markers have generally been found to decay faster in the environment than culturable *E. coli* and most enteric pathogens,^71,72^ and naturalized *E. coli* that grow in the environment in the absence of recent fecal contamination have been widely reported.^73^ A sanitation status index developed for MapSan study compounds was associated with soil *E. coli* counts,^20^ and the onsite sanitation intervention that we evaluated at the same households included in the present analysis reduced both the quantity of *E. coli* marker genes (but not *E. coli* colony counts) and the prevalence of enteric pathogens in soil.^21,24^ Improved sanitation and cement floors were associated with lower *E. coli* counts on household floors in peri-urban Peru,^74^ and community sanitation coverage was associated with lower *E. coli* across multiple environmental compartments in rural Bangladesh.^75^ Other evaluations of sanitation interventions have found limited impacts on *E. coli* in domestic environments,^11,76–78^ but water quality-related conditions have frequently been associated with *E. coli* concentrations in drinking water.^79–83^ Furthermore, increasing drinking water concentrations of *E. coli* or other fecal indicator bacteria were associated with elevated child diarrhea prevalence and reduced child linear growth in an individual participant data meta-analysis.^84^ Measuring drinking water quality prior to health outcome ascertainment is important for reliably characterizing the relationships between fecal markers and child health.^85,86^ A limitation of our analysis was the concurrent assessment of fecal markers in the environment and enteric pathogens in child stool; previous studies have reported relationships between fecal marker exposure and subsequent health outcomes that were not detectable in cross-sectional analyses of the same data.^85,87^

The clearest association we observed was between *E. coli* gene concentrations in household stored water and the prevalence of *Ascaris* in child stool. A relationship between drinking water quality and *Ascaris* was also observed in a large cluster-randomized trial in rural Kenya, where only the intervention arms that included water treatment reduced *Ascaris* prevalence in child stool.^88^ Although only significant for *Ascaris*, the prevalence of every pathogen we evaluated increased with higher concentrations of *E. coli* genes in household water. This broad pattern might reflect a general increase in child exposure to enteric pathogens through contaminated drinking water. The use of single measurements of household-level microbial water quality to represent child-level exposure to fecal contamination, as was the case in this analysis, has been shown to underestimate the relationship between fecal contamination and child enteric outcomes.^89^ We would expect the cross-sectional study design to further attenuate estimates and the modest sample size to limit their precision, suggesting that associations between impaired water quality and child pathogen exposure were likely underestimated.

That all pathogens were nevertheless detected more frequently in child stool when more *E. coli* was present in the household stored water suggests that *E. coli* genes indicated pathogen contamination of drinking water and increased probability that children in the household were exposed to enteric pathogens.

## Conclusions

We found that fecal markers in the domestic environment were not reliable predictors of child diarrhea nor detection of enteric pathogens in child stool. Human-associated marker HF183 in household stored water appeared to indicate lower risk of exposure to the most common pathogens, but this counter-intuitive association might be attributed to the limited sample size and infrequent detection of either human marker in water, casting further doubt on the suitability of existing human-associated fecal markers for exposure assessment in domestic environments. While *E. coli* colony counts in soil or water also were not associated with pathogen detection in stool, the concentration of *E. coli* gene markers in household stored water was positively associated with diarrhea and every pathogen. The association was significant for *Ascaris*, providing further evidence for a meaningful relationship between drinking water quality and *Ascaris* exposure for children,^88^ but weak and imprecise for all other outcomes.

Nevertheless, these more sensitive molecular *E. coli* measurements might prove informative about the risk of subsequent exposure to a variety of enteric pathogens through drinking water, particularly if implemented with prospective study designs and adequate sample sizes.^85,89^ Where molecular microbial detection in environmental samples is feasible, however, we advocate targeting enteric pathogens directly, in addition to or in lieu of molecular fecal markers. Although assessing the breadth of potentially relevant enteric pathogens remains costly,^68^ it generates more direct evidence of transmission and consistently demonstrated associations with both WASH conditions and child health in this setting.^69,70^

## Supporting Information

Stool processing and molecular analysis; environmental sample molecular analysis; fecal marker quantification and detection limits; laboratory quality control; multiple pathogen pooled regression model structure, interpretation, and prior distribution selection; posterior predicted probabilities for continuous exposures; fecal marker occurrence by outcome status; marginal prevalence estimates (PDF).

## Data Availability

The deidentified participant-level data and R code used in this analysis are available online at https://daholcomb.github.io/manuscripts/mapsan_mst/.

## Acknowledgment

We thank our implementing partner, Water and Sanitation for the Urban Poor; the data collection team at WE Consult; the laboratory staff at Instituto Nacional de Saúde; and Olimpio Zavale, Olivia Ginn, and Anna Stamatogiannakis. The table of contents graphic was created with BioRender. This study was funded by the United States Agency for International Development under Translating Research into Action, Cooperative Agreement No. GHS-A-00-09-00015-00, and was supported by the Bill & Melinda Gates Foundation (OPP1137224) and a National Institute of Environmental Health Sciences training grant (T32ES007018).

