## Supporting Information for "Associations between fecal contamination of the household environment and enteric pathogen detection in children living in Maputo, Mozambique"

### S1 Stool Processing and Molecular Analysis

Stool collection materials supplied to the caregivers of enrolled children included diapers and pre-labeled sterile sample bags. For older children who no longer wore diapers, we also provided a plastic potty and requested that the caregiver transfer stool collected in the potty to the provided diaper and place the diaper in the sterile sample bag for retrieval. When a stool sample was too liquid to aliquot directly into storage tubes, we instead stored a piece of the soaked diaper material in a 2 mL cryovial. Microscopy was not performed on the liquid stool samples. We isolated nucleic acids from bulk stool following the GPP manufacturer's protocol, pretreating bulk stool samples with 1 mL of ASL stool lysis buffer (Qiagen, Hilden, Germany). We eluted diaper samples in 2.5 mL of ASL buffer using a sterile 10 mL syringe to forcefully rinse the sample five times and eluted rectal swabs by agitating in 1 mL of lysis buffer for 1 min. Nucleic acids were isolated from 1 mL of pretreated stool or eluate using the QIAamp 96 Virus QIAcube HT Kit on the QIAcube HT platform (Qiagen).[1] We added the nonpathogenic RNA virus MS2 to the pretreated samples immediately before performing the extractions as a specimen processing control (SPC) to assess extraction efficiency and RT-PCR inhibition. Each set of extractions included at least one negative extraction control (NEC) containing only lysis buffer and MS2, as well as an extraction blank with lysis buffer only. Stool extracts were stored at 4 °C and analyzed by GPP within 24 hours. For approximately 10% of samples, we extracted and analyzed a second aliquot as a biological replicate. All GPP analyses included a no-template control (NTC) containing only molecular-grade water and PCR reagents.

### S2 Environmental Sample Molecular Analysis

We isolated DNA from soil sample filters using the DNeasy PowerSoil Kit (Qiagen) and from water sample filters with the DNA-EZ ST01 Kit (GeneRite, North Brunswick, NJ, USA).[2] Each extraction batch of up to 22 sample filters included both a positive control (PC), consisting of a clean filter spiked with $2 \times 10^8$ copies of composite DNA standard reference material (Table S1), and an NEC with a clean filter only.[3] We added 3 µg salmon testes DNA (Sigma-Aldrich, St. Louis, MO, USA) to each lysis tube (including PC and NEC tubes) as a specimen processing control.[4]

Table S1. Synthetic DNA reference material spiked into positive controls

| assays covered | sequence [5'-3'] | GenBank (base positions) | length [bases] |
|---|---|---|---|
| BacHum-UCD[5]<br>BacUni-UCD[5]<br>HF183/BacR287[6] | CCAGGATGGGATCATGAGTTCACATGTCCGCATGATTAA<br>AGGTATTTTCCGGTAGACGATGGGGATGCGTTCCATTAG<br>ATAGTAGGCGGGGTAACGGCCCACCTAGTCAACGATGGA<br>TAGGGGTTCTGAGAGGAAGGTCCCCCACATTGGAACTGA<br>GACACGGTCCAAACTCCTACGGGAGGCAGCAGTGAGGA<br>ATATTGGTCAATGGGCGATGGCCTGAACCAGCCAAGTAG<br>CGTGAAGGATGACTGCCCTATGGGTTGTAAACTTCTTTTA<br>TAAAGGAATAAAGTCGGGTATGCATACCCGTTTGCATGT<br>ACTTTATGAATAAGGATCGGCTAACTCCGTGCCAGCAGC<br>CGCGGTAATACGGAGGATCCGAGCGTTATCCGGATTTAT<br>TGGGTTTAAAGGGAGCGTAGATGGATGTTTAAGTCAGTT<br>GTGAAAGTTTGCGGCTCAACCGTAAAATTGCAGTTGATA<br>CTGGATGTCTTGAGTGCAGTTGAGGCAGGCGGAATTCGT<br>GGTGTAGCGGTGAAATGCTTAGATATCACGAAGAACTCC<br>GATTGCGAAGGCAGC | AB242142<br>(170-730) | 560 |
| GFD[7]<br>LA35[8] | TGGGTCTAATACCGGATACGACCATCTGCCGCATGGCGG<br>GTGGTGGAAAGTTTTTCGATTGGGGATGGGCTCGCGGCC<br>TATCAGTTTGTTGGTGGGGTAATGGCCTACCAAGGCGAC<br>GACGGGTAGCCGGCCTGAGAGGGCGACCGGCCACACTG<br>GGACTGAGACACGGCCCAGACTCCTACGGGAGGCAGCA<br>GTGGGGAATATTGCACAATGGGGGAAACCCTGATGCAGC<br>GACGCAGCGTGCGGGATGACGGCCTTCGGGTTGTAAACC<br>GCTTTCAGCAGGGAAGAAGCCTTCGGGTGACGGTACCTG<br>CAGAAGAAGTACCGGCTAACTACGTGCCAGCAGCCGCG<br>GTAATACGTAGGGTACGAGCGTTGTCCGGAATTATTGGG<br>CGTAAAGAGCTCGTAGGTGGTTGGTCACGTCTGCTGTGG<br>AAACGCAACGCTTAACGTTGCGCGGGCAGTGGGTACGGG<br>CTGACTAGAGTGCAGTAGGGGAGTCTGGAATTCCTGGTG<br>TAGCGGTGAAATGCGCAGATATCAGGAGGAACACCGGT<br>GGCGAAGGCGGGACTCTGGGCTGTGACTGACACTGGGG<br>AGCGAAAGCATTGCTAACAGTTcGGCTGAGCACTCTAGG<br>GAGACTGCCTTCGCAAGGAGGAGGAAGGTGAGGACGAC<br>GTCAAGTCATCATGGCCCTTACGCCTAGGGCTACACACG<br>TGCTACAATGGGATGTACAAAGAGACGCAATACCGCGA | FJ462358 (156-<br>746)<br>JN084061<br>(29-171) | 732 |
| EC23S857[9]<br>HAdV[10]<br>Mnif[11] | TAACTATGGTCATCGTTCGTCAGCAGTAACAGTAATTGCT<br>ACACCTGCTGAAACCACTGTCCCTTTTTCTTGGGCAACTC<br>TTGTTTATGTGTTGAAAGCGGAGGTCCTGAACCGGGTGT<br>TGGCTGTGCCGGACGTGGTGTAACAGTAGCTATGAAAAG<br>ACTTGAAAACTTAGGTGTTTTTGATAAGGATTTGGATGTA<br>GTCATTTATGGTGTACTTGGAGATGTTGTTTGCGGAGGTT<br>TTTCAGTGCCTTTACGTTCTCGGGCCAGGACGCCTCGGAG<br>TACCTGAGCCCCGGGCTGGTGCAGTTTGCCCGCGCCACC<br>GAGACGTACTTCAGCCTGAATAACAAGTTTAGAAACCCC<br>ACGGTGGCGCCTACGCATCTCCGGGGGTAGAGCACTGTT<br>TCGGCAAGGGGGTCATCCCGACTTACCAACCCGATGCAA<br>ACTGCGAATACCGGAGAATGTTATCACGGGAGACACACG<br>GCGGGT | AE015928<br>(4515891-<br>4515973)<br>AB019138<br>(192-363)<br>AC_000008<br>(18885-19000)<br>DQ682619<br>(847-954) | 475 |

We analyzed each environmental sample by three separate qPCR assays targeting fecal microbe genes and by the Sketa22 assay to assess PCR inhibition.[12,13] Approximately 10% of extracts from each sample type (stored water, household entrance soil, latrine entrance soil) were analyzed in duplicate. qPCR was performed on a CFX96 Touch thermocycler (Bio-Rad, Hercules, CA, USA) with an initial 10 minute, 95 °C incubation period and cycling conditions specified by the original developers for each assay (Table S2). The 25 µL reactions consisted of 12.5 µL TaqMan Environmental Master Mix 2.0 (Applied Biosystems, Waltham, MA, USA), 2.5 µL of 10x primers/probe mix, 5 µL molecular grade water, and 5 µL DNA template. Each instrument run included three NTCs and five-point, ten-fold dilution series from three extracted PCs to construct standard curves, corresponding to triplicate reactions with $10^5 – 10^1$ or $10^6 – 10^2$ target gene copies. Samples with Sketa22 quantification cycle (Cq) values > 3 above the mean Cq of extraction controls (both NEC and PC) were considered inhibited and diluted 1:5 for further analysis.

Table S2. qPCR assay cycling conditions

| assay[reference] | cycles | conditions | primer/probe (nM) | sequence (5'-3') |
|---|---|---|---|---|
| EC23S857[9] | 40 | 15 s: 95 °C<br>60 s: 60 °C | F (1000) | GGTAGAGCACTGTTTtGGCA |
| | | | R (1000) | TGTCTCCCGTGATAACtTTCTC |
| | | | P (80) | 6-FAM-TCATCCCGACTTACCAACCCG-BHQ1 |
| HF183/<br>BacR287[6] | 40 | 15 s: 95 °C<br>60 s: 60 °C | HF183 (1000) | ATCATGAGTTCACATGTCCG |
| | | | BacR287 (1000) | CTTCCTCTCAGAACCCCTATCC |
| | | | BacP234MGB (80) | 6-FAM-CTAATGGAACGCATCCC-BHQplus |
| Mnif [11] | 50 | 10 s: 95 °C<br>30 s: 57 °C | Mnif-202F (800) | GAAAGCGGAGGTCCTGAA |
| | | | Mnif-353R (800) | ACTGAAAAACCTCCGCAAAC |
| | | | Mnif-236P (240) | 6-FAM-CCGGACGTGGTGTAACAGTAGCTA-BHQ1 |
| Sketa22[12,13] | 40 | 15 s: 95 °C<br>60 s: 60 °C | SketaF2 (1000) | GGTTTCCGCAGCTGGG |
| | | | SketaR22 (1000) | CCGAGCCGTCCTGGTC |
| | | | SketaP2 (80) | 6-FAM-AGTCGCAGGCGGCCACCGT-BHQ1 |

### S3 Molecular Fecal Marker Quantification

Standard curves for quantifying molecular fecal markers were fit to ten-fold dilution series of positive controls that had been spiked with reference material for all three assays and extracted alongside each batch of samples.[3] Three gBlock linear DNA fragments (Integrated DNA Technologies, Skokie, Il, USA) containing composite reference sequences for the three fecal source tracking assays (as well as additional assays considered in the associated validation study) were used as standard reference material for the positive controls (Table S1).[3,14,15] Extracting the reference material accounted for loss of target DNA during extraction but induced greater variability between dilution series constructed from different PCs. We therefore allowed both the slopes and intercepts of the standard curves to vary by qPCR instrument run and extraction batch to account for the additional variation. We fit standard curves using Bayesian multilevel regression implemented in Stan through the *brms* package.[16–18] The resulting curves were relatively linear ($R^2 \geq 95\%$) when averaged across all instrument runs and extraction batches but were somewhat inefficient, particularly for HF183 (Table S3). We estimated fecal marker concentration as the mean of the posterior predictive distribution from the corresponding standard curve for the sample-specific extraction batch, instrument run, and measured Cq.

Table S3. Mean (95% CI) estimates of standard curve parameters

| **assay** | **intercept** | **slope** | **efficiency (%)** | **$R^2$** |
|---|---|---|---|---|
| EC23S857 | 47.9 (47.3, 48.6) | -3.50 (-3.64, -3.37) | 93.1 (88.1, 98.1) | 0.98 (0.97, 0.98) |
| HF183 | 47.5 (46.4, 48.6) | -3.85 (-4.07, -3.67) | 81.8 (76.1, 87.4) | 0.98 (0.97, 0.98) |
| Mnif | 48.8 (47.7, 50.3) | -3.47 (-3.79, -3.23) | 94.5 (83.6, 104.0) | 0.95 (0.93, 0.95) |

### S4 Detection Limits

The limit of detection (LOD) for each assay was determined using receiver operating characteristic (ROC) analysis.[19] We expressed assay LODs in terms of the cutoff Cq—the number of amplification cycles above which the target would be considered absent from the sample. We performed ROC analysis on the local validation study data reported in Holcomb et al. (2020), considering whole-Cq increments from 10 to the maximum number of cycles specified by the assay developers as the cutoff value.[3] Diagnostic sensitivity and specificity were calculated for each cutoff value, treating any reactions with a Cq value below the cutoff as positive. The highest Cq value that maximized the Youden index, $J = sensitivity + specificity - 1$, was considered the assay LOD (Table S4).[19,20]

Sample-specific process limits of detection (PLOD) were estimated for each sample from the assay LOD Cq values, the extraction batch- and instrument run-specific standard curve estimates, and the volume of water or equivalent dry mass of soil passed through the sample filter.[21,22] For *E. coli* enumerated by culture, we assumed an assay LOD of 1 colony forming unit (cfu) per plate and calculated the PLOD for each sample as the amount of sample represented by the least-diluted plate read for that sample.[3,23] This generally corresponded to PLODs of 1 cfu/100 mL water and 100 cfu/g wet soil, with varying sample-specific PLODs for soil by dry weight after accounting for moisture content. Because PLODs varied across samples, we present the mean (standard deviation) PLODs for each assay and sample matrix in Table S4.

Table S4. qPCR assay limits of detection and mean sample-specific process limits of detection

| | | process limit of detection | | | |
|---|---|---|---|---|---|
| | | soil $log_{10}$ copies/dry g | | water $log_{10}$ copies/100 mL | |
| **assay** | **assay LOD Cq** | **n** | **mean (SD)** | **n** | **mean (SD)** |
| EC23S857 | 39 | 241 | 4.51 (0.12) | 159 | 3.24 (0.13) |
| HF183 | 39 | 242 | 4.25 (0.33) | 159 | 2.84 (0.30) |
| Mnif | 41 | 241 | 4.18 (0.17) | 158 | 3.02 (0.10) |

### S5 Laboratory Quality Control

For approximately every 10 environmental samples, we also filtered sterile phosphate buffered saline (PBS) as a laboratory blank. All PBS filters were negative for culturable *E. coli* (n = 151). We included at least three NTC reactions and the NECs from the same extraction batches (typically three) as the samples on each qPCR plate. HF183 was not detected in any NTC (n = 46) or NEC (n = 46) reaction. Mnif was likewise absent from all NTC (n = 42) and NEC (n = 46) reactions. EC23S857 was detected at low concentrations in 2% of NTC (1/45) and 11% of NEC (5/46) reactions. The mean EC23S857 Cq in the negative control reactions in which it was detected was 38.1, with a minimum Cq of 37.1. These low concentrations only slightly exceeded the detection limit of 39 cycles and have been reported previously for molecular *E. coli* assays; the use of *E. coli* to produce the Environmental Master Mix is thought to occasionally result in residual *E. coli* DNA.[3,24] One soil sample had a Cq >3 above the mean Sketa22 Cq of the controls (positive and negative extraction) on the same plate and was diluted 1:5 in all further analyses to address PCR inhibition. Technical duplicate reactions were performed for a randomly selected 10% of samples of each sample type. The detection status agreed between duplicates for 94% (30/32) of EC23S857, 78% (25/32) of HF183, and 91% (29/32) of Mnif reaction pairs. We considered a sample positive if at least one of the duplicate reactions had a Cq lower than the detection limit for a given assay.

### S6 Multiple Pathogen Pooled Regression Model

Indices

samples: $i \in 1, \ldots, N$

compounds: $j \in 1, \ldots, J$

pathogens: $q \in 1, \ldots, Q$

covariates: $k \in 1, \ldots, K$

Structure

$$y_{i,j,q} \sim \text{Bernoulli}\left( p_{i,j,q}\right)$$

$$\text{logit}\left( p_{i,j,q}\right) = \alpha_{samp[i]} + \alpha_{comp[j]} + \alpha_{path[q]} + \beta_{path[q]} x_i + \gamma_{1,path[q]} z_{i,1} + \cdots + \gamma_{K,path[q]} z_{i,K}$$

$$\alpha_{samp[i]} \sim \text{Normal}\left(0,\ \sigma_{samp}\right)$$

$$\alpha_{comp[j]} \sim \text{Normal}\left(0,\ \sigma_{comp}\right)$$

$$\begin{bmatrix} \alpha_{path[q]} \\ \beta_{path[q]} \\ \gamma_{1,path[q]} \\ \vdots \\ \gamma_{K,path[q]} \end{bmatrix} \sim \text{MVN}\left( \begin{bmatrix} \alpha \\ \beta \\ \gamma_1 \\ \vdots \\ \gamma_K \end{bmatrix}, \Sigma \right) \quad \text{(S1)}$$

$$\Sigma = \tau \times \Omega \times \tau$$

$$\tau = \begin{pmatrix} \sigma_\alpha & 0 & \cdots & & 0 \\ 0 & \sigma_\beta & & & \vdots \\ \vdots & & \sigma_{\gamma_1} & & \\ & & & \ddots & 0 \\ 0 & \cdots & & 0 & \sigma_{\gamma_K} \end{pmatrix}$$

$$\Omega = \begin{pmatrix} 1 & \rho_{\alpha,\beta} & \cdots & \rho_{\alpha,\gamma_K} \\ \rho_{\beta,\alpha} & 1 & & \vdots \\ \vdots & & & \\ & & \ddots & \\ \rho_{\gamma_K,\alpha} & \cdots & & 1 \end{pmatrix}$$

Interpretation

Table S5. Interpretation of binary pooled outcome model terms

| Term | Interpretation |
|---|---|
| $y_{i,j,q}$ | Detection status of pathogen $q$ in stool $i$ from compound $j$ |
| $p_{i,j,q}$ | Probability of detecting pathogen $q$ in stool $i$ from compound $j$ |
| $\alpha_{samp[i]}$ | Sample-specific random effect for sample $i$; the change in baseline log-odds from the overall mean for all pathogens in sample $i$ |
| $\sigma_{samp}$ | Group standard deviation for sample-level random effects; larger value indicates greater clustering of pathogen outcomes by sample and results in less pooling of information between samples |
| $\alpha_{comp[j]}$ | Compound-specific random effect for compound $j$ |
| $\sigma_{comp}$ | Group standard deviation of compound-level random effects; larger value indicates greater clustering of pathogen outcomes by compound and results in less pooling of information between compounds |
| $\alpha_{path[q]}$ | Pathogen-specific intercept for pathogen $q$; the log-odds of pathogen $q$ for an unexposed child in the reference group for all covariates |
| $\alpha$ | Group mean of the intercept across all $Q$ pathogens; the log-odds of a generic pathogen for an unexposed child in the reference group for all covariates |
| $\beta_{path[q]}$ | Pathogen-specific effect of exposure on the log-odds of pathogen $q$, conditional on the other covariates; exp $(\beta_{path[q]})$ gives the conditional odds ratio for the effect of exposure on pathogen $q$ |
| $x_i$ | Exposure status for sample $i$ |
| $\beta$ | Group mean conditional exposure effect across the population of $Q$ pathogens; the pooled estimate of the conditional effect of exposure on a generic enteric pathogen |
| $\gamma_{k,path[q]}$ | Conditional effect of covariate $k$ on the log-odds of pathogen $q$ |
| $z_{i,k}$ | Value of covariate $k$ for sample $i$ |
| $\gamma_k$ | Group mean conditional effect of covariate $k$ across the population of $Q$ pathogens |
| $\Sigma$ | Symmetric $K + 2$ matrix with the group variance of each pathogen-varying effect ($\alpha$, $\beta$, $\gamma_k$) on the diagonal and their covariances on the off-diagonals; decomposes into the scale matrix $\tau$ and the correlation matrix $\Omega$ |
| $\tau$ | The scale matrix for pathogen-varying effects $\alpha, \beta, \gamma_k$: a $K + 2$ diagonal matrix of the group standard deviations $\sigma_\alpha, \sigma_\beta, \sigma_{\gamma_k}$; these standard deviations reflect the extent to which each effect varies across the group of $Q$ pathogens, with larger values indicating greater differences between pathogens |
| $\Omega$ | The correlation matrix for pathogen-varying effects $\alpha, \beta, \gamma_k$: a $K + 2$ square matrix with the pairwise correlations $\rho_{\alpha,\beta}$, etc. on the off-diagonals; these correlations reflect how the different effects co-vary by pathogen. The correlations between $\alpha$ and each of the other effects are of particular interest as they indicate how low or high background pathogen prevalence may modulate the exposure effect and the other covariate effects |

### S7 Prior Distributions

We specified prior distributions for the mixed effects logistic regression models as described in the data analysis plan for the long-term follow-up study to the MapSan trial.[25] Weakly informative prior distributions were used to regularize parameter estimates and aid computation. Because all parameters except for the correlation matrix Ω were estimated on the log-odds scale, we specified standard normal priors for all slope parameters and standard half-normal priors for all group-level standard deviation parameters:

$$
\begin{aligned}
&\alpha \sim \mathrm{Normal}(-1, 2), \qquad \sigma_{\alpha} \sim \mathrm{Normal}^{+}(0,1) \\
&\beta \sim \mathrm{Normal}(0,1), \qquad \sigma_{\beta} \sim \mathrm{Normal}^{+}(0,1) \\
&\gamma_k \sim \mathrm{Normal}(0,1), \qquad \sigma_{\gamma_k} \sim \mathrm{Normal}^{+}(0,1) \\
&\sigma_{samp} \sim \mathrm{Normal}^{+}(0, 1), \\
&\sigma_{comp} \sim \mathrm{Normal}^{+}(0, 1) \\
&\Omega \sim \mathrm{LKJcorr}(2)
\end{aligned}
\tag{S2}
$$

The $\mathrm{Normal}(-1,2)$ prior on the overall intercept parameter $\alpha$, which is on the log-odds scale, implies an expected prevalence for a generic pathogen of $\mathrm{logit}^{-1}(-1) \approx 27\%$. This corresponds to a 68% chance that the prevalence falls between 5% and 73% and a 95% chance that the prevalence is between 0.7% and 95%.[26,27] Combined with a standard half-normal (positive constrained) hyperprior on the group standard deviation $\sigma_{\alpha}$ for additional variation, the pathogen-specific intercept $\alpha_{path[q]}$ for each pathogen $q$ is capable of taking any prevalence value supported by the data with gentle regularization towards prevalence between 0% and 50%, reflecting prior knowledge of the typical frequency of individual pathogens in this setting. Such regularization can facilitate better computational behavior by softly constraining the sampler away from infeasible values to explore the parameter space more efficiently. By encouraging light smoothing, regularization can also help stabilize parameter estimates; unconstrained estimation, by contrast, can be susceptible to undue influence by ordinary noise in the data, particularly in the context of complex, high-dimensional models and with the relatively small datasets typical of environmental microbial assessments.[27,28]

Similarly, the standard normal priors for $\beta$ and $\gamma_k$ on the log-odds scale imply that each covariate has a 95% chance of altering the prevalence of a pathogen with the expected value for $\alpha_{path[q]}$ (corresponding to a background prevalence of 27%) by up to 22 percentage points. Representing a large but feasible potential effect size, these priors likewise provide gentle regularization towards more moderate parameter values, centered on no effect, while allowing sufficiently strong evidence in the data to produce substantial effect estimates in either direction. In addition to assisting computationally, this mild regularization towards the null helps control false discoveries that would be expected to arise from ordinary noise in the data when comparing multiple outcomes by introducing light smoothing that requires stronger patterns in the data to register as credible effects.[29,30] The degree of smoothing applied to effect estimates for individual pathogens, $\beta_{path[q]}$ and $\gamma_{k,path[q]}$, is learned from the data and represented by the group standard deviations $\sigma_\beta$ and $\sigma_{\gamma_k}$. Larger group standard deviation estimates correspond to greater variability by pathogen and thus impose relatively less smoothing on the effect estimates. The independent standard half-normal priors on the group standard deviations likewise allows for a wide range of plausible values when considered on the probability scale, while discouraging extremely large values.[26,31,32]

The group correlations between the pathogen varying effects ($\rho_{\alpha,\beta}$, $\rho_{\beta,\gamma_1}$, etc.) were collectively given an LKJ prior on the correlation matrix, which imposes the [-1,1] constraint on correlations but otherwise assigns approximately uniform prior density across that range when shape parameter $\eta = 1$.[28,33] Larger values of $\eta$ increase the weight around zero, which can be used to regularize the correlation estimates towards smaller absolute values. We selected $\eta = 2$ to provide mild regularization for computational purposes while still permitting strong correlations when warranted.[28,34]

### S8 Posterior Predicted Probabilities for Continuous Exposures

Unlike posterior predicted probabilities for binary exposure variables, where unexposed status is indicated by a value of 0 and exposed status represented by 1, the change in probability for a one-unit increase in the value of a continuous exposure variable is also dependent on the initial value of the exposure variable for each observation in the model. Simply comparing the predicted probability when each exposure value is incremented by one unit with the predicted probability for the original exposure does not account for the non-linear relationship with exposure on the probability scale. We instead estimated the instantaneous slope (i.e., first derivative) on the probability scale at the observed exposure value of each observation in the model.[35–37] We used finite differences to numerically approximate the instantaneous slope, which corresponds to the prevalence difference (PD) as the expected change in pathogen prevalence for a $\log_{10}$ increase in *E. coli* concentration.[38] For each stool sample, we predicted posterior probabilities after subtracting a small constant $(5 \times 10^{-5})$ from the observed *E. coli* concentration, and again after adding $5 \times 10^{-5}$ to the observed concentration. The instantaneous slope for observation $i$ was calculated by subtracting the predicted probability for the lower concentration from the predicted probability at the higher concentration, then dividing this difference in predicted probabilities by $10^{-4}$ (the difference in concentration between the two prediction scenarios):

$$slope_i = \frac{P\left(x_i + \frac{\epsilon}{2}\right) - P\left(x_i - \frac{\epsilon}{2}\right)}{\epsilon}, \qquad \epsilon = 1 \times 10^{-4} \tag{S3}$$

where $x_i$ is the observed concentration. We then estimated the marginal prevalence ratio (PR) by adding the estimated slope to the posterior predicted probability for the original observed concentration $x_i$ before dividing by the predicted probability at the observed concentration:

$$PR_i = \frac{P(x_i) + slope_i}{p(x_i)} \tag{S4}$$

### S9 Fecal Marker Occurrence by Outcome Status

The mean *E. coli* concentration and human marker detection frequency among environmental samples paired with pathogen-positive and pathogen-negative child stool are reported in Table S6 for household stored water and Table S7 for domestic soil.

Table S6. Fecal marker occurrence in household stored water samples by outcome detection status

| | | Household stored water | | | | | |
|---|---|---|---|---|---|---|---|
| | | ***E. coli* colonies** $\log_{10}$ **cfu/100 mL** | | ***E. coli* genes** $\log_{10}$ **copies/100 mL** | | **HF183 detection** | |
| **Outcome** | **Status** | **N** | **mean (SD)** | **N** | **mean (SD)** | **N** | **n (%)** |
| Any bacteria | Negative | 58 | 1.4 (1.4) | 59 | 4.3 (0.7) | 59 | 13 (22) |
| | Positive | 127 | 1.4 (1.3) | 127 | 4.2 (0.8) | 127 | 17 (13) |
| *C. difficile* | Negative | 175 | 1.4 (1.3) | 176 | 4.3 (0.8) | 176 | 28 (16) |
| | Positive | 10 | 1.6 (1.6) | 10 | 4.1 (0.8) | 10 | 2 (20) |
| *Campylobacter* | Negative | 162 | 1.4 (1.4) | 163 | 4.2 (0.8) | 163 | 24 (15) |
| | Positive | 23 | 1.8 (1.2) | 23 | 4.3 (0.6) | 23 | 6 (26) |
| ETEC | Negative | 136 | 1.4 (1.4) | 137 | 4.2 (0.8) | 137 | 24 (18) |
| | Positive | 49 | 1.5 (1.3) | 49 | 4.3 (0.8) | 49 | 6 (12) |
| STEC | Negative | 177 | 1.4 (1.4) | 178 | 4.2 (0.8) | 178 | 29 (16) |
| | Positive | 8 | 1.9 (1.0) | 8 | 4.6 (0.7) | 8 | 1 (12) |
| *Shigella* | Negative | 90 | 1.5 (1.4) | 92 | 4.3 (0.7) | 92 | 23 (25) |
| | Positive | 95 | 1.3 (1.3) | 94 | 4.2 (0.8) | 94 | 7 (7) |
| Any viruses | Negative | 156 | 1.5 (1.3) | 156 | 4.3 (0.8) | 156 | 24 (15) |
| | Positive | 29 | 1.2 (1.5) | 30 | 4.2 (0.8) | 30 | 6 (20) |
| Norovirus | Negative | 159 | 1.5 (1.3) | 159 | 4.3 (0.8) | 159 | 25 (16) |
| | Positive | 26 | 1.2 (1.5) | 27 | 4.2 (0.8) | 27 | 5 (19) |
| Any protozoa | Negative | 86 | 1.5 (1.4) | 87 | 4.2 (0.8) | 87 | 20 (23) |
| | Positive | 99 | 1.4 (1.3) | 99 | 4.3 (0.7) | 99 | 10 (10) |
| *Cryptosporidium* | Negative | 178 | 1.4 (1.4) | 179 | 4.2 (0.8) | 179 | 28 (16) |
| | Positive | 7 | 1.8 (1.3) | 7 | 4.6 (0.7) | 7 | 2 (29) |
| *Giardia* | Negative | 93 | 1.5 (1.4) | 94 | 4.2 (0.8) | 94 | 22 (23) |
| | Positive | 92 | 1.3 (1.3) | 92 | 4.3 (0.7) | 92 | 8 (9) |
| Any STH | Negative | 82 | 1.4 (1.4) | 84 | 4.2 (0.8) | 84 | 17 (20) |
| | Positive | 81 | 1.5 (1.4) | 79 | 4.3 (0.8) | 79 | 11 (14) |
| *Ascaris* | Negative | 132 | 1.3 (1.4) | 132 | 4.2 (0.8) | 132 | 22 (17) |
| | Positive | 31 | 1.9 (1.2) | 31 | 4.7 (0.4) | 31 | 6 (19) |
| *Trichuris* | Negative | 92 | 1.5 (1.3) | 94 | 4.2 (0.7) | 94 | 20 (21) |
| | Positive | 71 | 1.4 (1.4) | 69 | 4.3 (0.8) | 69 | 8 (12) |
| Diarrhea | Negative | 168 | 1.5 (1.4) | 168 | 4.2 (0.8) | 168 | 28 (17) |
| | Positive | 18 | 1.1 (0.9) | 19 | 4.6 (0.8) | 19 | 2 (11) |

Table S7. Fecal marker occurrence in domestic soil samples by outcome detection status

| | | Domestic soil | | | | | | | |
|---|---|---|---|---|---|---|---|---|---|
| | | ***E. coli*** **colonies log$_{10}$ cfu/dry g** | | ***E. coli*** **genes log$_{10}$ copies/dry g** | | **HF183 detection** | | **Mnif detection** | |
| **Outcome** | **Status** | **N** | **mean (SD)** | **N** | **mean (SD)** | **N** | **n (%)** | **N** | **n (%)** |
| Any bacteria | Negative | 105 | 3.8 (1.1) | 107 | 6.7 (1.0) | 107 | 49 (46) | 106 | 43 (41) |
| | Positive | 228 | 3.9 (1.0) | 242 | 6.8 (1.1) | 242 | 116 (48) | 241 | 114 (47) |
| *C. difficile* | Negative | 320 | 3.8 (1.0) | 330 | 6.8 (1.0) | 330 | 161 (49) | 328 | 147 (45) |
| | Positive | 13 | 3.8 (1.2) | 19 | 6.8 (1.0) | 19 | 4 (21) | 19 | 10 (53) |
| *Campylobacter* | Negative | 291 | 3.9 (1.0) | 307 | 6.8 (1.0) | 307 | 136 (44) | 305 | 133 (44) |
| | Positive | 42 | 3.8 (1.0) | 42 | 7.0 (1.0) | 42 | 29 (69) | 42 | 24 (57) |
| ETEC | Negative | 245 | 3.8 (1.1) | 256 | 6.8 (1.1) | 256 | 119 (46) | 254 | 110 (43) |
| | Positive | 88 | 4.0 (1.0) | 93 | 6.9 (1.0) | 93 | 46 (49) | 93 | 47 (51) |
| STEC | Negative | 317 | 3.9 (1.0) | 333 | 6.8 (1.1) | 333 | 158 (47) | 331 | 148 (45) |
| | Positive | 16 | 3.8 (0.9) | 16 | 6.8 (0.9) | 16 | 7 (44) | 16 | 9 (56) |
| *Shigella* | Negative | 161 | 3.8 (1.1) | 168 | 6.8 (1.0) | 168 | 78 (46) | 167 | 68 (41) |
| | Positive | 172 | 3.9 (1.0) | 181 | 6.9 (1.1) | 181 | 87 (48) | 180 | 89 (49) |
| Any viruses | Negative | 280 | 3.9 (1.0) | 296 | 6.8 (1.1) | 296 | 140 (47) | 295 | 134 (45) |
| | Positive | 53 | 3.6 (1.2) | 53 | 6.7 (1.0) | 53 | 25 (47) | 52 | 23 (44) |
| Norovirus | Negative | 285 | 3.9 (1.0) | 301 | 6.8 (1.1) | 301 | 145 (48) | 300 | 138 (46) |
| | Positive | 48 | 3.6 (1.2) | 48 | 6.6 (1.0) | 48 | 20 (42) | 47 | 19 (40) |
| Any protozoa | Negative | 157 | 3.8 (1.1) | 163 | 6.8 (1.1) | 163 | 83 (51) | 163 | 66 (40) |
| | Positive | 176 | 3.9 (0.9) | 186 | 6.9 (1.0) | 186 | 82 (44) | 184 | 91 (49) |
| *Cryptosporidium* | Negative | 323 | 3.8 (1.0) | 336 | 6.8 (1.0) | 336 | 162 (48) | 334 | 150 (45) |
| | Positive | 10 | 4.2 (0.9) | 13 | 6.4 (1.4) | 13 | 3 (23) | 13 | 7 (54) |
| *Giardia* | Negative | 167 | 3.8 (1.1) | 176 | 6.7 (1.1) | 176 | 87 (49) | 176 | 74 (42) |
| | Positive | 166 | 3.9 (0.9) | 173 | 6.9 (1.0) | 173 | 78 (45) | 171 | 83 (49) |
| Any STH | Negative | 142 | 3.9 (1.0) | 152 | 6.7 (1.0) | 152 | 68 (45) | 151 | 71 (47) |
| | Positive | 153 | 3.8 (1.0) | 159 | 6.9 (1.1) | 159 | 78 (49) | 159 | 71 (45) |
| *Ascaris* | Negative | 231 | 3.8 (1.0) | 247 | 6.7 (1.0) | 247 | 109 (44) | 246 | 113 (46) |
| | Positive | 64 | 3.9 (1.2) | 64 | 7.1 (1.0) | 64 | 37 (58) | 64 | 29 (45) |
| *Trichuris* | Negative | 161 | 3.9 (1.0) | 171 | 6.8 (1.0) | 171 | 82 (48) | 170 | 77 (45) |
| | Positive | 134 | 3.8 (1.1) | 140 | 6.9 (1.1) | 140 | 64 (46) | 140 | 65 (46) |
| Diarrhea | Negative | 303 | 3.8 (1.0) | 319 | 6.8 (1.0) | 319 | 145 (45) | 317 | 143 (45) |
| | Positive | 32 | 4.3 (0.8) | 32 | 7.3 (0.9) | 32 | 19 (59) | 32 | 14 (44) |

### S10 Marginal Prevalence Estimates

We used posterior prediction to estimate the marginal prevalence distribution of each outcome (individual pathogen, pathogen class, diarrhea) under three fecal indicator exposure scenarios: 1) the observed distribution of exposures among all participant observation-environmental sample pairs; 2) all observation-sample pairs set to unexposed; and 3) all observation-sample pairs set to exposed. Prevalence differences and prevalence ratios were calculated from the posterior prevalence predictions as described in the main text and above. The “observed” exposure scenario reflected the empirical prevalence of each outcome among the participant observation-sample pairs included in the model for a given outcome, environmental sample matrix, and fecal indicator; because the number of such pairs varied, the prevalence estimates for the observed scenario were similar but not identical to the crude prevalence for each outcome when considering only the participant observations reported in Table 2 of the main text. The “unexposed” and “exposed” scenarios were only evaluated for the human fecal indicator presence/absence for which a binary exposed/unexposed status was meaningful. Marginal PD estimates for diarrhea, individual pathogens, and pooled across all pathogens are displayed in Figure S1. The constraint at zero absolute prevalence produced truncated 95% CIs on the PD for some lower prevalence pathogens, for which the PR intervals on the relative scale were more symmetrical (Figure 1). Exposure associations with composite pathogen class variables are presented as marginal PR estimates in Figure S2 and as marginal PD estimates in Figure S3. The posterior predicted prevalence under each exposure scenario (observed, unexposed, exposed) and the corresponding marginal PR and PD estimates are reported in Table S8.

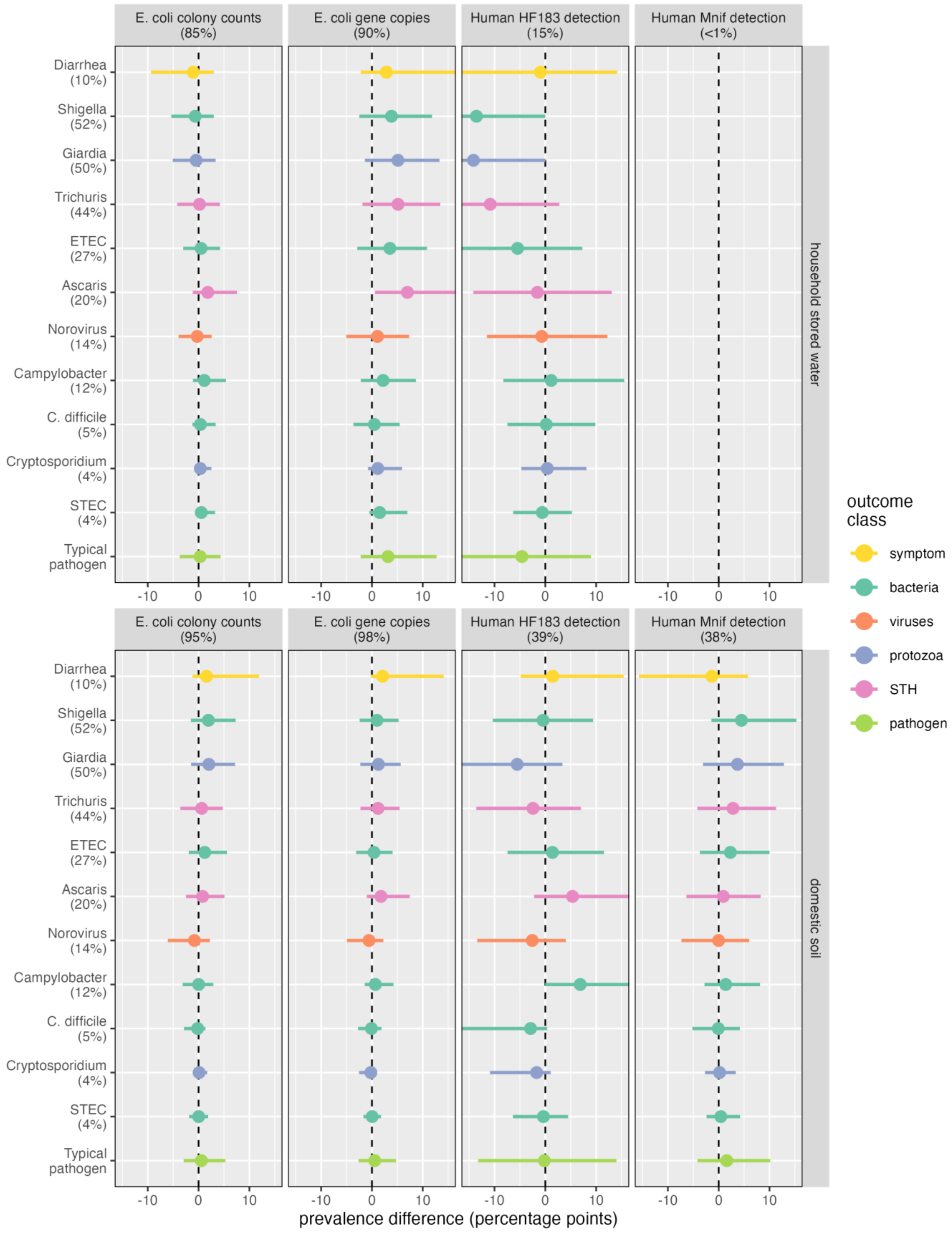

Figure S1. Mean and 95% CI marginal prevalence difference estimates of diarrhea and of enteric pathogens in stool for a ten-fold increase in E. coli concentration or detection of a human fecal marker.

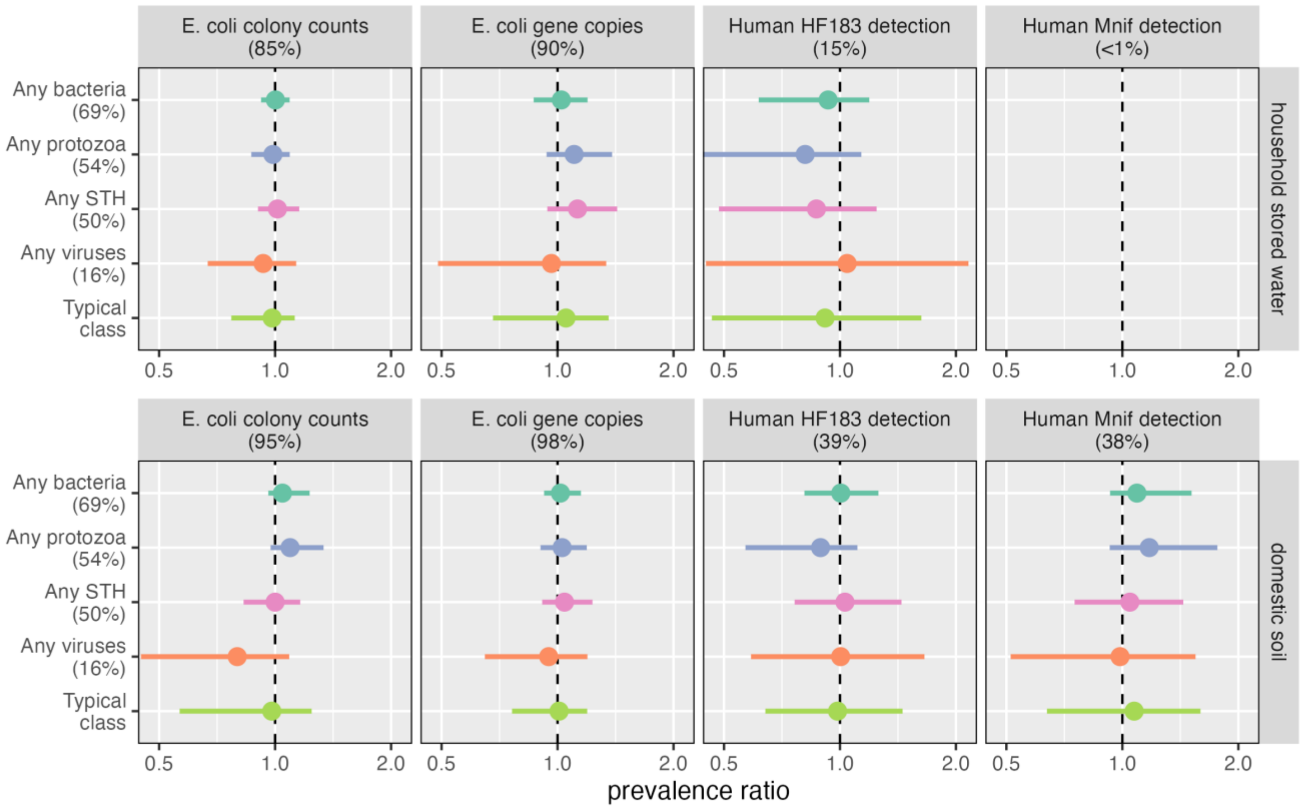

Figure S2. Mean and 95% CI marginal prevalence ratio estimates of detecting any pathogen of a given class for a ten-fold increase in *E. coli* concentration or detection of a human fecal marker.

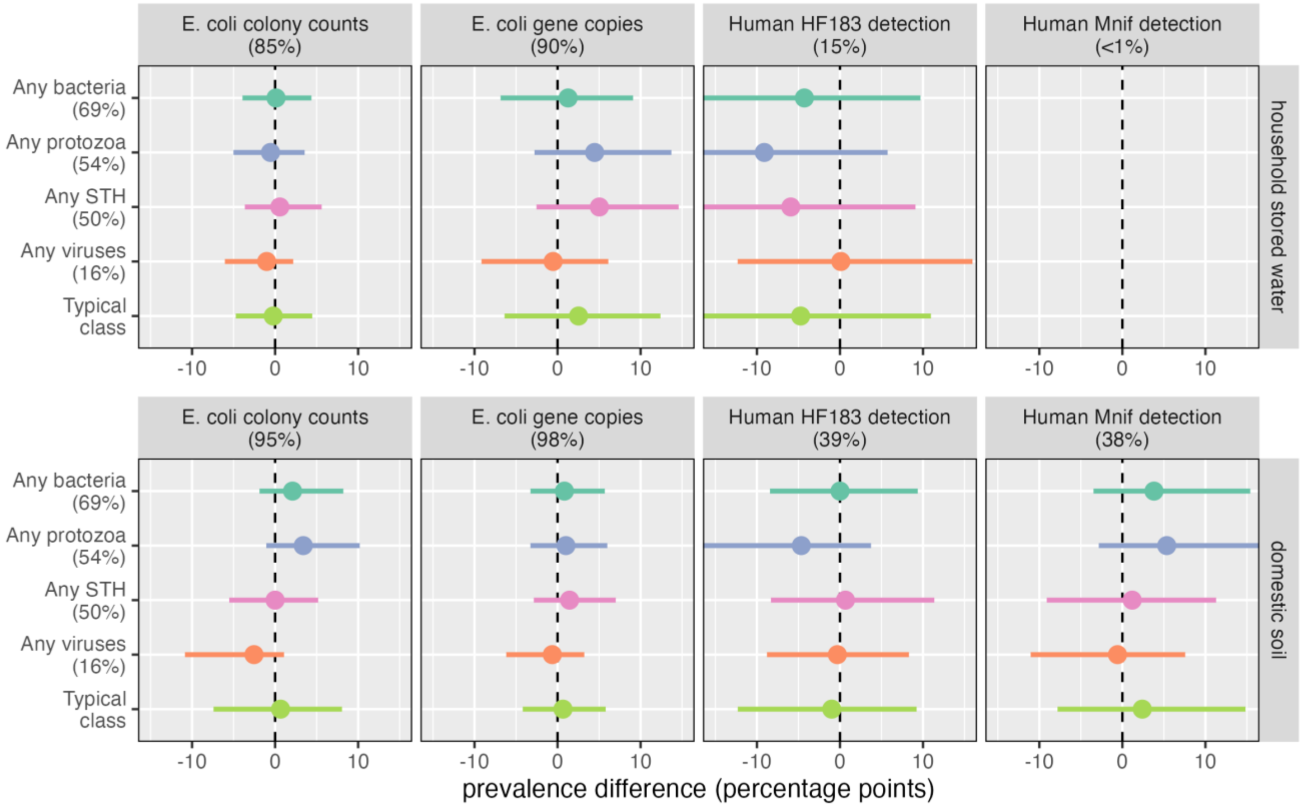

Figure S3. Mean and 95% CI marginal prevalence difference estimates of detecting any pathogen of a given class for a ten-fold increase in *E. coli* concentration or detection of a human fecal marker.

Table S8. Mean (95% CI) estimates of posterior predicted prevalence, prevalence differences, and prevalence ratios for each outcome

| Outcome | Exposure | Posterior predicted prevalence[a] | | | PD | PR |
|---|---|---|---|---|---|---|
| | | Observed % | Unexposed % | Exposed % | pp[b] | |
| ***Stored water samples*** | | | | | | |
| Any bacteria | *E. coli* colonies | 68 (25, 96) | | | 0.1 (-3.9, 4.4) | 1.0 (0.9, 1.1) |
| | *E. coli* genes | 68 (25, 96) | | | 1.3 (-6.9, 9.1) | 1.0 (0.9, 1.2) |
| | HF183 | 68 (25, 96) | 69 (26, 96) | 65 (21, 95) | -4.3 (-20.7, 9.7) | 0.9 (0.6, 1.2) |
| *C. difficile* | *E. coli* colonies | 6 (0, 28) | | | 0.4 (-1.2, 3.4) | 1.1 (0.8, 1.4) |
| | *E. coli* genes | 6 (0, 28) | | | 0.5 (-3.6, 5.5) | 1.1 (0.4, 1.6) |
| | HF183 | 6 (0, 28) | 6 (0, 27) | 6 (0, 30) | 0.2 (-7.5, 9.8) | 1.1 (0.4, 2.8) |
| *Campylobacter* | *E. coli* colonies | 12 (2, 36) | | | 1.1 (-1.1, 5.4) | 1.1 (0.9, 1.3) |
| | *E. coli* genes | 12 (2, 35) | | | 2.2 (-2.2, 8.7) | 1.2 (0.8, 1.5) |
| | HF183 | 12 (2, 35) | 12 (2, 34) | 13 (2, 40) | 1.2 (-8.3, 15.5) | 1.1 (0.5, 2.4) |
| ETEC | *E. coli* colonies | 26 (5, 59) | | | 0.5 (-3.0, 4.2) | 1.0 (0.9, 1.2) |
| | *E. coli* genes | 26 (6, 59) | | | 3.5 (-2.9, 10.8) | 1.2 (0.9, 1.4) |
| | HF183 | 26 (6, 60) | 27 (6, 61) | 22 (4, 55) | -5.4 (-19.8, 7.3) | 0.8 (0.4, 1.3) |
| STEC | *E. coli* colonies | 5 (0, 18) | | | 0.5 (-0.7, 3.2) | 1.1 (0.9, 1.5) |
| | *E. coli* genes | 5 (0, 18) | | | 1.5 (-0.5, 7.0) | 1.3 (0.9, 1.9) |
| | HF183 | 5 (1, 17) | 5 (1, 17) | 5 (0, 17) | -0.6 (-6.3, 5.2) | 0.9 (0.3, 2.2) |
| *Shigella* | *E. coli* colonies | 51 (7, 95) | | | -0.7 (-5.4, 3.0) | 1.0 (0.8, 1.1) |
| | *E. coli* genes | 50 (7, 95) | | | 3.8 (-2.5, 11.8) | 1.1 (0.9, 1.4) |
| | HF183 | 51 (6, 95) | 53 (8, 95) | 39 (3, 91) | -13.5 (-35.6, 0.0) | 0.7 (0.3, 1.0) |
| Any viruses | *E. coli* colonies | 16 (2, 47) | | | -1.0 (-6.0, 2.2) | 0.9 (0.7, 1.1) |
| | *E. coli* genes | 17 (2, 46) | | | -0.5 (-9.2, 6.1) | 1.0 (0.5, 1.3) |
| | HF183 | 17 (2, 47) | 17 (2, 47) | 17 (2, 50) | 0.1 (-12.3, 16.0) | 1.0 (0.4, 2.2) |
| Norovirus | *E. coli* colonies | 14 (2, 40) | | | -0.3 (-3.9, 2.6) | 1.0 (0.8, 1.2) |
| | *E. coli* genes | 14 (2, 40) | | | 1.1 (-5.0, 7.3) | 1.1 (0.7, 1.4) |
| | HF183 | 14 (2, 40) | 15 (2, 40) | 14 (2, 42) | -0.7 (-11.5, 12.2) | 1.0 (0.4, 2.0) |
| Any protozoa | *E. coli* colonies | 53 (13, 90) | | | -0.6 (-5.0, 3.6) | 1.0 (0.9, 1.1) |
| | *E. coli* genes | 53 (13, 90) | | | 4.5 (-2.8, 13.7) | 1.1 (0.9, 1.4) |
| | HF183 | 53 (12, 91) | 54 (15, 91) | 45 (9, 87) | -9.1 (-28.2, 5.7) | 0.8 (0.4, 1.1) |
| *Cryptosporidium* | *E. coli* colonies | 4 (0, 18) | | | 0.3 (-0.8, 2.5) | 1.1 (0.8, 1.4) |
| | *E. coli* genes | 4 (0, 17) | | | 1.1 (-0.8, 5.9) | 1.3 (0.8, 1.9) |
| | HF183 | 4 (0, 17) | 4 (0, 17) | 5 (0, 20) | 0.4 (-4.7, 8.1) | 1.2 (0.4, 3.1) |
| *Giardia* | *E. coli* colonies | 49 (10, 89) | | | -0.5 (-5.1, 3.4) | 1.0 (0.9, 1.1) |
| | *E. coli* genes | 49 (10, 90) | | | 5.1 (-1.4, 13.3) | 1.1 (1.0, 1.4) |
| | HF183 | 49 (8, 90) | 51 (11, 90) | 37 (5, 83) | -14.1 (-34.5, 0.0) | 0.7 (0.3, 1.0) |
| Any STH | *E. coli* colonies | 49 (12, 87) | | | 0.6 (-3.6, 5.6) | 1.0 (0.9, 1.2) |
| | *E. coli* genes | 48 (12, 86) | | | 5.0 (-2.5, 14.6) | 1.1 (0.9, 1.4) |
| | HF183 | 48 (11, 86) | 49 (12, 87) | 43 (9, 84) | -5.9 (-22.9, 9.1) | 0.9 (0.5, 1.2) |

| Outcome | Exposure | Posterior predicted prevalence[a] Observed % | Unexposed % | Exposed % | PD pp[b] | PR |
|---|---|---|---|---|---|---|
| *Ascaris* | *E. coli* colonies | 19 (3, 50) | | | 1.8 (-1.1, 7.5) | 1.1 (0.9, 1.4) |
| | *E. coli* genes | 19 (2, 54) | | | 7.0 (0.6, 21.2) | 1.4 (1.1, 2.0) |
| | HF183 | 19 (3, 51) | 20 (3, 51) | 18 (3, 51) | -1.6 (-14.2, 13.1) | 0.9 (0.4, 1.8) |
| *Trichuris* | *E. coli* colonies | 43 (8, 84) | | | 0.2 (-4.2, 4.2) | 1.0 (0.9, 1.1) |
| | *E. coli* genes | 42 (8, 83) | | | 5.1 (-1.9, 13.4) | 1.2 (0.9, 1.4) |
| | HF183 | 42 (8, 83) | 43 (9, 84) | 33 (5, 77) | -10.8 (-28.7, 2.8) | 0.7 (0.3, 1.1) |
| Typical pathogen[c] | *E. coli* colonies | 23 (0, 84) | | | 0.3 (-3.7, 4.3) | 1.0 (0.8, 1.3) |
| | *E. coli* genes | 23 (0, 84) | | | 3.2 (-2.2, 12.7) | 1.2 (0.8, 1.7) |
| | HF183 | 23 (0, 84) | 24 (0, 84) | 19 (0, 74) | -4.6 (-26.6, 9.0) | 0.9 (0.4, 2.1) |
| Diarrhea | *E. coli* colonies | 10 (0, 68) | | | -1.1 (-9.4, 3.0) | 0.8 (0.3, 1.3) |
| | *E. coli* genes | 11 (0, 69) | | | 2.8 (-2.2, 18.7) | 1.4 (0.7, 2.2) |
| | HF183 | 11 (0, 67) | 11 (0, 68) | 10 (0, 67) | -1.0 (-18.7, 14.2) | 1.0 (0.2, 3.0) |
| ***Soil samples*** | | | | | | |
| Any bacteria | *E. coli* colonies | 68 (10, 99) | | | 2.1 (-1.9, 8.2) | 1.0 (1.0, 1.2) |
| | *E. coli* genes | 69 (11, 99) | | | 0.8 (-3.3, 5.7) | 1.0 (0.9, 1.1) |
| | HF183 | 69 (11, 99) | 69 (11, 99) | 69 (11, 99) | 0.0 (-8.4, 9.4) | 1.0 (0.8, 1.3) |
| | Mnif | 69 (11, 99) | 67 (10, 99) | 71 (12, 99) | 3.8 (-3.5, 15.4) | 1.1 (0.9, 1.5) |
| *C. difficile* | *E. coli* colonies | 4 (0, 26) | | | -0.2 (-2.9, 1.4) | 0.9 (0.6, 1.2) |
| | *E. coli* genes | 5 (0, 35) | | | -0.1 (-2.7, 1.9) | 1.0 (0.7, 1.2) |
| | HF183 | 6 (0, 37) | 7 (0, 42) | 4 (0, 27) | -2.9 (-19.6, 0.4) | 0.6 (0.2, 1.2) |
| | Mnif | 6 (0, 35) | 6 (0, 36) | 5 (0, 35) | -0.1 (-5.2, 4.2) | 1.0 (0.5, 1.6) |
| *Campylobacter* | *E. coli* colonies | 13 (1, 44) | | | 0.0 (-3.1, 2.9) | 1.0 (0.8, 1.2) |
| | *E. coli* genes | 12 (1, 43) | | | 0.7 (-1.4, 4.2) | 1.1 (0.9, 1.3) |
| | HF183 | 12 (0, 45) | 9 (0, 35) | 16 (1, 53) | 6.9 (0.0, 24.4) | 2.1 (1.0, 4.1) |
| | Mnif | 12 (1, 43) | 11 (1, 40) | 13 (1, 44) | 1.4 (-2.8, 8.1) | 1.2 (0.8, 1.7) |
| ETEC | *E. coli* colonies | 26 (2, 70) | | | 1.2 (-1.9, 5.6) | 1.1 (0.9, 1.2) |
| | *E. coli* genes | 27 (2, 70) | | | 0.4 (-3.1, 4.1) | 1.0 (0.9, 1.2) |
| | HF183 | 27 (2, 70) | 26 (2, 69) | 27 (2, 71) | 1.4 (-7.4, 11.5) | 1.1 (0.7, 1.6) |
| | Mnif | 26 (2, 70) | 25 (2, 68) | 28 (3, 71) | 2.3 (-3.7, 10.0) | 1.1 (0.9, 1.5) |
| STEC | *E. coli* colonies | 5 (0, 23) | | | 0.0 (-1.9, 1.9) | 1.0 (0.7, 1.3) |
| | *E. coli* genes | 5 (0, 22) | | | 0.0 (-1.7, 1.8) | 1.0 (0.7, 1.2) |
| | HF183 | 5 (0, 22) | 5 (0, 23) | 5 (0, 21) | -0.4 (-6.4, 4.5) | 1.0 (0.4, 1.9) |
| | Mnif | 5 (0, 22) | 5 (0, 21) | 5 (0, 23) | 0.4 (-2.4, 4.2) | 1.1 (0.7, 1.7) |
| *Shigella* | *E. coli* colonies | 52 (3, 97) | | | 1.9 (-1.5, 7.3) | 1.1 (1.0, 1.3) |
| | *E. coli* genes | 52 (3, 97) | | | 1.0 (-2.4, 5.2) | 1.0 (0.9, 1.2) |
| | HF183 | 52 (3, 97) | 52 (3, 97) | 51 (3, 97) | -0.4 (-10.4, 9.4) | 1.0 (0.7, 1.4) |
| | Mnif | 52 (3, 97) | 50 (3, 97) | 54 (4, 98) | 4.5 (-1.4, 15.3) | 1.2 (1.0, 1.7) |

| Outcome | Exposure | Posterior predicted prevalence[a] | | | PD | PR |
|---|---|---|---|---|---|---|
| | | Observed % | Unexposed % | Exposed % | pp[b] | |
| Any viruses | *E. coli* colonies | 16 (0, 65) | | | -2.5 (-10.9, 1.1) | 0.8 (0.4, 1.1) |
| | *E. coli* genes | 16 (0, 62) | | | -0.6 (-6.2, 3.2) | 0.9 (0.6, 1.2) |
| | HF183 | 15 (0, 61) | 16 (0, 61) | 15 (0, 61) | -0.3 (-8.8, 8.3) | 1.0 (0.6, 1.7) |
| | Mnif | 15 (0, 60) | 16 (0, 61) | 15 (0, 60) | -0.6 (-11.0, 7.6) | 1.0 (0.5, 1.5) |
| Norovirus | *E. coli* colonies | 14 (1, 49) | | | -0.8 (-6.1, 2.2) | 0.9 (0.6, 1.1) |
| | *E. coli* genes | 14 (1, 47) | | | -0.6 (-4.9, 2.2) | 1.0 (0.7, 1.1) |
| | HF183 | 14 (1, 47) | 15 (1, 49) | 12 (1, 44) | -2.6 (-13.4, 4.0) | 0.8 (0.5, 1.3) |
| | Mnif | 14 (1, 46) | 14 (1, 46) | 14 (1, 46) | 0.0 (-7.3, 6.0) | 1.0 (0.6, 1.5) |
| Any protozoa | *E. coli* colonies | 53 (4, 97) | | | 3.4 (-1.1, 10.2) | 1.1 (1.0, 1.3) |
| | *E. coli* genes | 53 (4, 97) | | | 1.0 (-3.3, 6.0) | 1.0 (0.9, 1.2) |
| | HF183 | 53 (4, 97) | 55 (5, 97) | 51 (4, 96) | -4.7 (-17.5, 3.7) | 0.9 (0.6, 1.1) |
| | Mnif | 53 (4, 97) | 50 (4, 96) | 56 (5, 97) | 5.3 (-2.8, 17.4) | 1.2 (0.9, 1.8) |
| *Cryptosporidium* | *E. coli* colonies | 3 (0, 17) | | | 0.1 (-1.1, 1.7) | 1.0 (0.7, 1.3) |
| | *E. coli* genes | 4 (0, 21) | | | -0.2 (-2.5, 1.0) | 0.9 (0.6, 1.2) |
| | HF183 | 4 (0, 22) | 5 (0, 24) | 3 (0, 17) | -1.7 (-10.9, 1.0) | 0.7 (0.2, 1.4) |
| | Mnif | 4 (0, 21) | 4 (0, 21) | 4 (0, 21) | 0.2 (-2.7, 3.3) | 1.1 (0.6, 1.7) |
| *Giardia* | *E. coli* colonies | 50 (5, 93) | | | 2.0 (-1.5, 7.2) | 1.1 (1.0, 1.2) |
| | *E. coli* genes | 49 (5, 94) | | | 1.3 (-2.3, 5.7) | 1.0 (0.9, 1.2) |
| | HF183 | 49 (5, 94) | 52 (5, 94) | 46 (4, 93) | -5.5 (-17.1, 3.4) | 0.9 (0.6, 1.1) |
| | Mnif | 49 (4, 94) | 47 (4, 93) | 51 (5, 94) | 3.7 (-3.1, 12.8) | 1.1 (0.9, 1.5) |
| Any STH | *E. coli* colonies | 52 (4, 96) | | | 0.0 (-5.5, 5.2) | 1.0 (0.8, 1.2) |
| | *E. coli* genes | 51 (3, 96) | | | 1.4 (-2.9, 7.0) | 1.0 (0.9, 1.2) |
| | HF183 | 51 (4, 96) | 51 (3, 96) | 52 (4, 97) | 0.7 (-8.3, 11.4) | 1.0 (0.8, 1.4) |
| | Mnif | 51 (4, 97) | 51 (4, 96) | 52 (4, 97) | 1.2 (-9.1, 11.3) | 1.0 (0.8, 1.4) |
| *Ascaris* | *E. coli* colonies | 22 (1, 67) | | | 0.8 (-2.5, 5.1) | 1.0 (0.9, 1.2) |
| | *E. coli* genes | 21 (1, 66) | | | 1.8 (-1.0, 7.4) | 1.1 (0.9, 1.4) |
| | HF183 | 21 (1, 66) | 18 (1, 61) | 24 (2, 70) | 5.4 (-2.2, 19.1) | 1.4 (0.9, 2.4) |
| | Mnif | 21 (1, 66) | 20 (1, 65) | 21 (1, 66) | 0.9 (-6.4, 8.2) | 1.1 (0.7, 1.5) |
| *Trichuris* | *E. coli* colonies | 45 (4, 91) | | | 0.6 (-3.6, 4.8) | 1.0 (0.9, 1.2) |
| | *E. coli* genes | 45 (4, 92) | | | 1.2 (-2.3, 5.4) | 1.0 (0.9, 1.2) |
| | HF183 | 45 (4, 92) | 46 (4, 92) | 43 (4, 91) | -2.4 (-13.6, 7.0) | 0.9 (0.6, 1.3) |
| | Mnif | 45 (4, 92) | 44 (4, 91) | 46 (4, 93) | 2.8 (-4.2, 11.3) | 1.1 (0.9, 1.5) |
| Typical pathogen | *E. coli* colonies | 23 (0, 88) | | | 0.6 (-2.9, 5.3) | 1.0 (0.7, 1.2) |
| | *E. coli* genes | 24 (0, 89) | | | 0.5 (-2.6, 4.7) | 1.0 (0.8, 1.2) |
| | HF183 | 24 (0, 89) | 24 (0, 90) | 23 (0, 88) | -0.2 (-13.2, 14.0) | 1.0 (0.3, 2.6) |
| | Mnif | 23 (0, 89) | 22 (0, 87) | 24 (0, 90) | 1.6 (-4.2, 10.2) | 1.1 (0.7, 1.6) |

| Outcome | Exposure | Posterior predicted prevalence[a] | | | PD | PR |
|---|---|---|---|---|---|---|
| | | Observed % | Unexposed % | Exposed % | pp[b] | |
| Diarrhea | *E. coli* colonies | 10 (0, 75) | | | 1.6 (-1.2, 11.9) | 1.3 (0.8, 1.8) |
| | *E. coli* genes | 9 (0, 74) | | | 2.1 (-0.2, 14.1) | 1.4 (0.9, 1.9) |
| | HF183 | 9 (0, 74) | 9 (0, 70) | 10 (0, 76) | 1.5 (-4.8, 15.4) | 1.4 (0.6, 3.2) |
| | Mnif | 9 (0, 74) | 10 (0, 77) | 9 (0, 71) | -1.3 (-15.7, 5.8) | 0.9 (0.3, 2.0) |

[a] Predicted population-level prevalence of outcome assuming: 1) the observed exposure values; 2) the entire population was unexposed; and 3) the entire population was exposed. Unexposed and exposed predicted prevalence only apply to the binary exposures assessed for human fecal markers

[b] pp: percentage points

[c] Pooled exposure effect across all the pathogens included in the model; predicted using the group means and standard deviations estimated for the pathogen-varying slopes and intercepts

(12) Haugland, R. A.; Siefring, S. C.; Wymer, L. J.; Brenner, K. P.; Dufour, A. P. Comparison of Enterococcus Measurements in Freshwater at Two Recreational Beaches by Quantitative

Polymerase Chain Reaction and Membrane Filter Culture Analysis. *Water Res* **2005**, *39* (4), 559–568. https://doi.org/10.1016/j.watres.2004.11.011.

(25) Holcomb, D.; Capone, C.; Cumbane, V.; Knee, J.; Kowaslky, E.; Lai, A.; et al. *Data Analysis Plan: Long-Term Impact of an Urban Sanitation Intervention on Child Health in Low-Income Neighborhoods of Maputo City, Mozambique: A Cross-Sectional Follow-up Five Years Post-*

*Intervention in the Maputo Sanitation (MapSan) Trial. Version 4 (Revised 2023-01-20).*; 2023. https://osf.io/e7pvk/.

(26) Holcomb, D. A.; Knee, J.; Capone, D.; Sumner, T.; Adriano, Z.; Nalá, R.; et al. Impacts of an Urban Sanitation Intervention on Fecal Indicators and the Prevalence of Human Fecal Contamination in Mozambique. *Environ Sci Technol* **2021**, *55* (17), 11667–11679. https://doi.org/10.1021/acs.est.1c01538.

(27) Gelman, A.; Carpenter, B. Bayesian Analysis of Tests with Unknown Specificity and Sensitivity. *J R Stat Soc C: Appl Stat* **2020**, *69* (5), 1269–1283. https://doi.org/10.1111/rssc.12435.

(28) McElreath, R. *Statistical Rethinking: A Bayesian Course with Examples in R and Stan*, 2nd ed.; Chapman & Hall/CRC Texts in Statistical Science; CRC Press: Boca Raton, 2020.
